# Airborne Transmission of COVID-19 and Mitigation Using Box Fan Air Cleaners in a Poorly Ventilated Classroom

**DOI:** 10.1101/2021.03.11.21253395

**Authors:** Ruichen He, Wanjiao Liu, John Elson, Rainer Vogt, Clay Maranville, Jiarong Hong

## Abstract

Many indoor places, including aged classrooms and offices, prisons, homeless shelters, etc., are poorly ventilated but resource-limited to afford expensive ventilation upgrade or commercial air purification systems, raising concerns on the safety of opening activities in these places in the era of COVID-19 pandemic. To address this challenge, using computational fluid dynamics, we conducted a systematic investigation of airborne transmission in a classroom equipped with a single horizontal unit ventilator (HUV) and evaluate the performance of low-cost box fan air cleaner for risk mitigation. Our study shows that placing box fan air cleaners in the classroom results in a substantial reduction of airborne transmission risk across the entire space. The air cleaner can achieve optimal performance when placed near the asymptomatic patient. However, without knowing the location of the patient, the performance of the cleaner is optimal near the HUV with the air flowing downwards. In addition, we find that it is more efficient in reducing aerosol concentration and spread in the classroom by adding air cleaners in comparison with raising the flow rate of HUV alone. The number and placement of air cleaners need to be adjusted to maintain its efficacy for larger classrooms and to account for the thermal gradient associated with human thermal plume and hot ventilation air during cold seasons. Overall, our study shows that box fan air cleaners can serve as an effective low-cost alternative for mitigating airborne transmission risks in poorly ventilated spaces.

## I. INTRODUCTION

Increasing evidence has shown that airborne transmission is an important pathway that leads to the spread of COVID-19^1–4^. Compared to outdoor settings, the risk of airborne transmission is significantly higher for various congregated indoor activities^5–7^. Improved ventilation has been commonly recommended as an important preventive measure to reduce the risk of indoor airborne transmission ^8^. By replacing contaminated air with clean air, ventilation can help lower the concentration of particulate matters and reduce the probability of exposure to virus-containing aerosols^9,10^. Particularly, one study has shown that a low infection probability of less than 1% can be achieved with a ventilation rate above typical recommended values^11^. However, many indoor spaces are poorly ventilated, including a large number of old public-school classrooms^11,12^ and offices^13^. These classrooms are especially prone to higher risks of airborne transmission, due to aged infrastructure, high population density, and extended hours of operation that can lead to high levels of aerosol accumulation. Studies have shown that opening window is an effective approach to alleviate aerosol accumulation ^14^, but is hard to implement during cold/hot seasons, and in many classrooms with no operable windows. Another suggested mitigation approach is to upgrade the existing central HVAC system ^15^, but the high cost impedes its implementation in resource-limited indoor spaces.

As an alternative approach, portable air purifiers are broadly used for risk mitigation in these poorly ventilated spaces^16^. It has been recently demonstrated in a classroom with no ventilation that the usage of HEPA grade purifiers can significantly reduce the aerosol concentration level ^17^. Nevertheless, the commercial purifiers used for public spaces such as classrooms and shared offices typically require a clean air delivery rate (CADR) of larger than 400 cfm^18^ with price ranging from $ 400 to above $4000^19^. Such high costs limit the wide adoption of this mitigation approach, particularly in resource-limited indoor places^10,14^, including public schools, prisons, down-market offices, shelters, life care centers, etc. To cope with this challenge, a low-cost air cleaner, constructed using readily available air filter panels and a box fan was proposed^20^. Unlike its commercial counterparts, the performance of this low-cost system had not previously been evaluated in a systematic fashion.

Therefore, using computational fluid dynamics (CFD) approach, our current study aims to provide a systematic assessment of using these low-cost air cleaners as an alternative approach for risk mitigation in poorly ventilated indoor spaces. Since the outbreak of SARS in 2003, CFD has been employed as an effective tool to assess airborne transmission risks under various indoor and outdoor settings^21–25^. Particularly, Lin et al.^26^ first simulated airborne transmission due to coughing in a well-ventilated classroom with 12 air exchange per hour (ACH) under different ventilation designs and showed that mixing ventilation leads to the highest aerosol concentration compared to displacement and stratum ventilation designs. Using a classroom of similar size (under 7.5 ACH), Zhang et al.^27^ investigated transmission caused by continuous talking and demonstrated the superiority of displacement over mixing ventilation in lowering aerosol concentration and spread. Abuhegazy et al.^28^ systematically evaluated the effect of the location of an asymptomatic individual (referred to as infector hereafter) and the size of particles generated by the infector on airborne transmission in a well-ventilated classroom (8.6 ACH) with distributed ventilation. They showed a substantial fraction (24% to 50%) of particles can be removed by the ventilation and opening the window can further increases the fraction of removal to 69%. In contrast, in a classroom with a single site ventilation, Shao et al. ^29^ showed that ventilation can only extract a small fraction of aerosols (∼3%) even under an exceedingly high ventilation (i.e., 30 ACH) with majority of aerosols depositing on surfaces due to the presence of stable flow circulation regions in the space.

Despite these past efforts, very few studies focused on airborne transmission in poorly ventilated classrooms. These classrooms are widely present in public schools^30^. They are usually equipped with a single horizontal unit ventilator (HUV, unit ventilator is the most common type of HVAC system used in public schools) operating at air exchange rate of around 2 ACH, significantly lower than the ventilation used in abovementioned investigations. Furthermore, as pointed out earlier, no study has systematically evaluated these low-cost air cleaners including the influence of placement and design on their performance, although some have studied the location effect of the air cleaner under other settings^31,32^.

To fill in this gap, our study focuses on investigating the airborne transmission under poorly ventilated classroom settings and the efficacy of the corresponding mitigation strategies using low-cost box fan air cleaners. The work is conducted using CFD with air cleaner models characterized using experiments. The present paper is structured as follows: Sec II describes the design of our simulation including the design and characterization of low-cost air cleaner model and the setups of different simulation cases. Successively, in Sec III, we present our simulation results evaluating the influence of the placement and flow direction of air cleaners, room size as well as the thermal gradient in the air on the air cleaner performance. The results are also compared with those from the simulation cases using only enhanced ventilation (no air cleaner placed in the room). Finally, the conclusions and discussions are provided in Sec IV.

## II. METHODOLOGY

### A. Box fan air cleaner model

The low-cost box fan air cleaner used in the study is designed and constructed by Ford^33^. As shown in Figure 1a and b, the air cleaner is comprised of an easy-to-assemble die-cut cardboard support, a box fan of 0.5 × 0.5 m ^2^ cross section, and a 0.5 × 0.5 × 0.1 m ^3^ air filter with a standard minimum efficiency reporting value (MERV) of 13. The filter panel is placed inside the folded base with the fan placed on top. The fan operates on high for maximum filtration, discharging clean air downward as it pulls in unfiltered air from above.

**FIG. 1.**
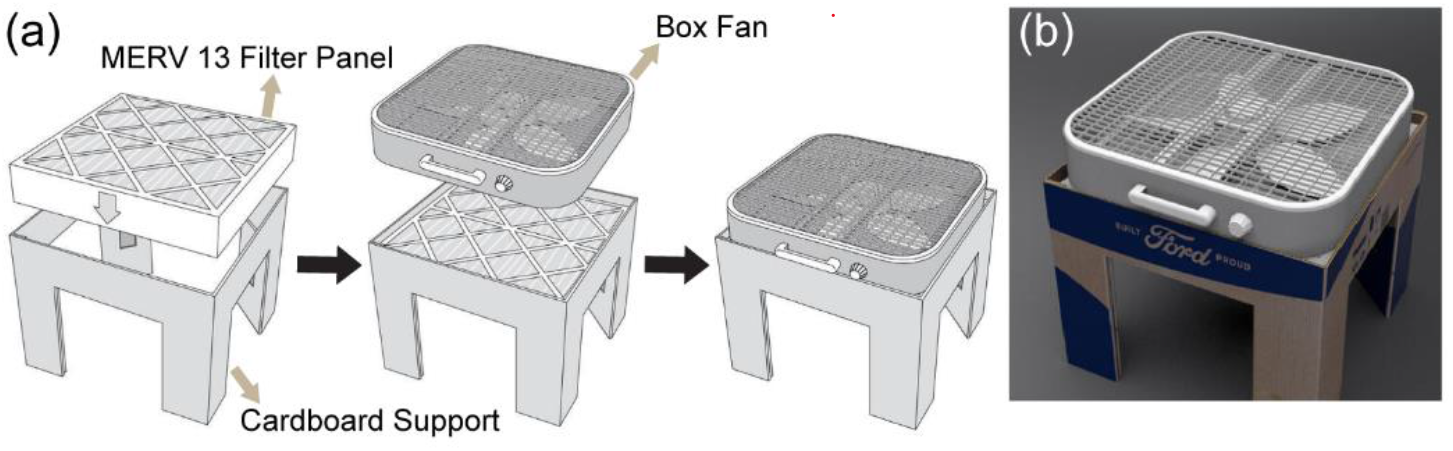
(a) Schematic showing the composition and assembly procedure of the box fan air cleaner designed by Ford. (b) Photo of the box fan air cleaner.

To characterize the flow rate and inlet velocity profile of the box fan air cleaner, a vane anemometer (OMEGA) is used to measure the velocity at 35 locations distributed at the inlet surface of the box fan air cleaner (Figure 2a). The inlet velocity profile is found to be nearly uniform over 80% of the total area in the center as shown in Figure 2b, with an average velocity of 1.5 m/s with a standard deviation of 0.2 m/s. Based on these measurements, the total flow rate is calculated to be about 0.2 m^3^/s. Accordingly, for simplification, we use a flow rate of 0.2 m^3^/s and a uniform inlet velocity profile for the air cleaner model in the simulations. To evaluate the filtration performance of the filter panel used in the box-fan air cleaner, the commercially available air filter panel (Tri -Pleat Green 20204SP, Tri-Dim Mann & Hummel) performance is measured using the ASHRAE 52.2-2017 test standard^34^ at an independent test lab using KCl as the challenge aerosol, which reported performance exceeding the MERV 13 performance standard. Therefore, in the simulation, we set the filtration efficiency of the air cleaner to be 100% for simplification. To assess the CADR of the air cleaner, two experiments are conducted at two independent laboratories following the ANSI/AHAM AC-1 test standard^35^ using tobacco smoke as the challenge particle. Tobacco smoke CADR is reported as 213 cfm (362 m ^3^/hr.) in the first lab and 231 cfm (392 m^3^/hr.) at the second lab. The difference in performance is likely due to setup and measurement differences.

**FIG. 2.**
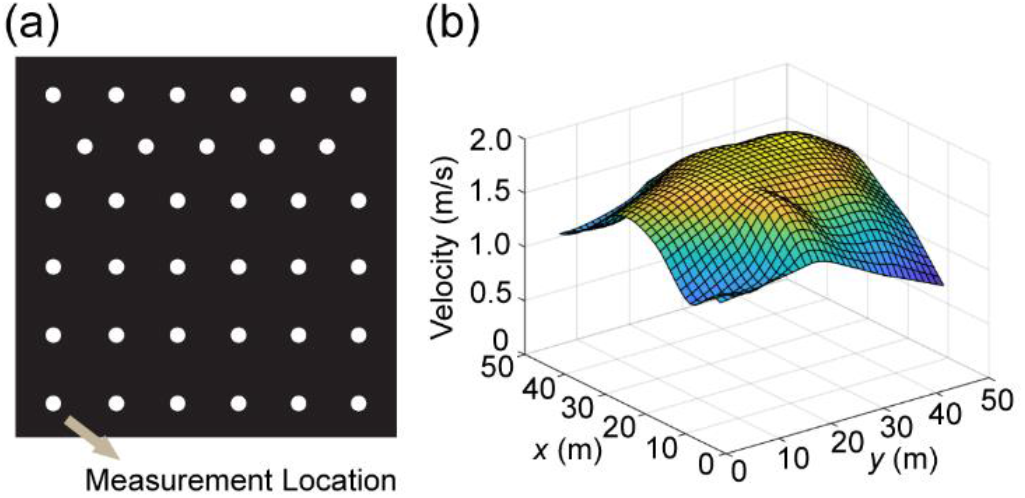
(a) Schematic showing the locations of anemometer measurements used to characterize the inflow conditions of the box fan air cleaner. (b) Inlet velocity profile of the air cleaner obtained from the anemometer measurements.

### B. Numerical simulation

CFD simulation is conducted using OpenFOAM-2012 platform, with the Eulerian -Lagrangian framework for simulating gas-particle phases. The implicit unsteady shear stress transport *k*-*ω* turbulence model is used with low Reynolds number modification to model the flow turbulence, which has been used in previous studies investigating the aerosol dispersion from human respiratory activities ^36–38^. Air flow is calculated using a compressible solver to model the buoyancy forces based on the following equations:

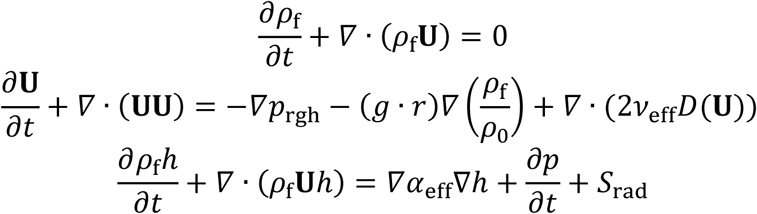

In the equations, *ρ*_f_ is fluid density, *U* represents the flow velocity, *g* is the gravity acceleration, *p*_rgh_ is the new defined pressure under the assumption of boussinesq approximation, *r* is the position vector, *v*_eff_ is the kinemetric viscosity, *h* is the enthalpy, *τ* is the stress tensor, *α*_eff_ represents the effective thermal diffusivity, and *S*_rad_ is the source term for the radiative heat transfer. To handle the convective terms, the second-order upwind scheme is implemented. For the diffusion terms, the Gauss-linear second order approach is used. For the coupling of the pressure and the velocity, the Pressure-Implicit with Splitting of Operator (PISO) algorithm is applied. The minimum residuals for the convergence of pressure, and velocity are 10^−8^, and 10^−12^, respectively.

As for the particle movement simulation, one -way coupled Eulerian-Lagrangian approach is applied to predict the deposition and dispersion of each particle. Particles are assumed to be spherical and particle -particle interactions are ignored. The particle motion is tracked by using the time integration of Newton’s second law. The translational motion of each particle is governed by the Maxey -Riley equation. To determine the particle velocity *u*_*i*P_, and position *x*_*i*P_, such equation is solved for each particle, which is given by:

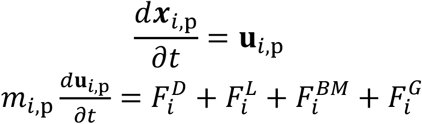

In the equations, *i* is the particle ID, *u*_p_ is the particle velocity, *m*_p_ is the particle mass, *F*^D^ represents the drag force, *F*^L^ is the lift force, *F*^G^ is the gravitational force, and *F*^BM^ is the force induced by Brownian motion. The drag force uses the following form:

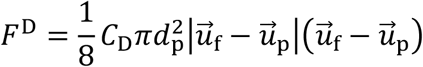

In the equation, *d*_p_ is the particle diameter, and *u*_f_ is the velocity of the fluid. The drag coefficient, *C*_D_, is determined by the following equation:

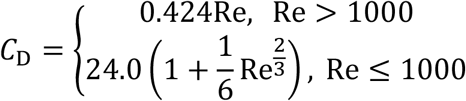

The lift force is of the form:

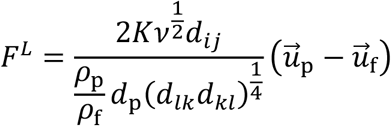

In the equation, *K* = 2.594 is the constant coefficient of Saffman’s lift force, *v* is the kinematic viscosity, and *ρ*_p_ is the particle density. The density of water is used for *ρ*_p_, as the particles are mostly water^39^. The deformation rate tensor, *d*_*ij*_, is defined as

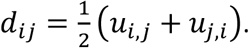

The Brownian motion induced force is of the following forms:

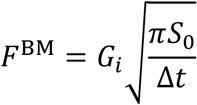

with *G*_*i*_ are the zero-mean, unit variance independent Gaussian random numbers, Δ*t* is the time step used in the simulation, and

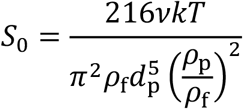

In the equation, *k* = 1.38 × 10^−23^J/K is the Boltzmann constant and *T* is the absolute temperature of the fluid. In the end, the combine effect of gravity and buoyancy is

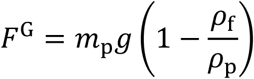

There are various forms of particle interactions. Only particle-wall interactions such as deposit and rebound are considered in the simulation. The standard wall interaction functions provided by OpenFOAM are implemented in the study to simulate the interaction between the particle and the wall patch, which has been validated in previous study in comparison with experiment data^40^, and has been used for different types of simulation studies^41^.

Table 1 summarizes all the simulation cases presented in the current study including (i) baseline cases (Cases A and Case B), (ii) cases to evaluate the placement effect of air cleaner on their performance (Cases A1 to Case A4, Case B1 to Case B4, and Case A12 and Case B12), (iii) cases to evaluate the inflow direction of air cleaners on their performance (Cases FA2 and Case FB2), (iv) cases with only enhanced ventilation for comparison with the cases using air cleaners (Cases VA and Case VB), (v) cases to evaluate the effect of room size on air cleaners performance (Cases LB, Case LB2, Case LB12, and Case LB22), and (vi) cases that include the thermal effect (Cases TA2, Case TB2, Case TA2H, and Case TB2H). Specifically, for baseline cases, the computational domain is selected to simulate a classroom of 10 × 5 × 3 m^3^, as shown in Figure 3a. The classroom is equipped with a horizontal unit ventilator (HUV) simplified as a 0.3 × 0.8 × 1.5 m^3^ cuboid placed next to the wall (Figure 3b). The inlet and outlet dimensions of HUV are 1.2 × 0.5 m^2^ and 1.2 × 1 m^2^, respectively. An outlet pressure boundary condition is applied at the HUV inlet patch while a constant mass flow rate boundary condition is used for the outlet. The flow rate of the HVU is set as 325 cfm (0.15 m^3^/s, corresponding to 2 ACH for the simulated classroom size) with 50% filtration efficiency, which is used to simplify adding same amount of outside clean air with the recycled polluted air. The room air temperature is set as 24 °C. Zero gradient temperature and no-slip wall boundary conditions are applied to all the wall in the domain. An asymptomatic instructor, referred to as the infector hereafter, are placed in the front (Location A) or the middle (Location B) of the classroom. The simulations are conducted over a 50-minute duration with continuous particle injection at 110 particles per second^42^ with a mean diameter of 2 μm representing an asymptomatic instructor giving a 50-minute lecture.

**TABLE 1.**
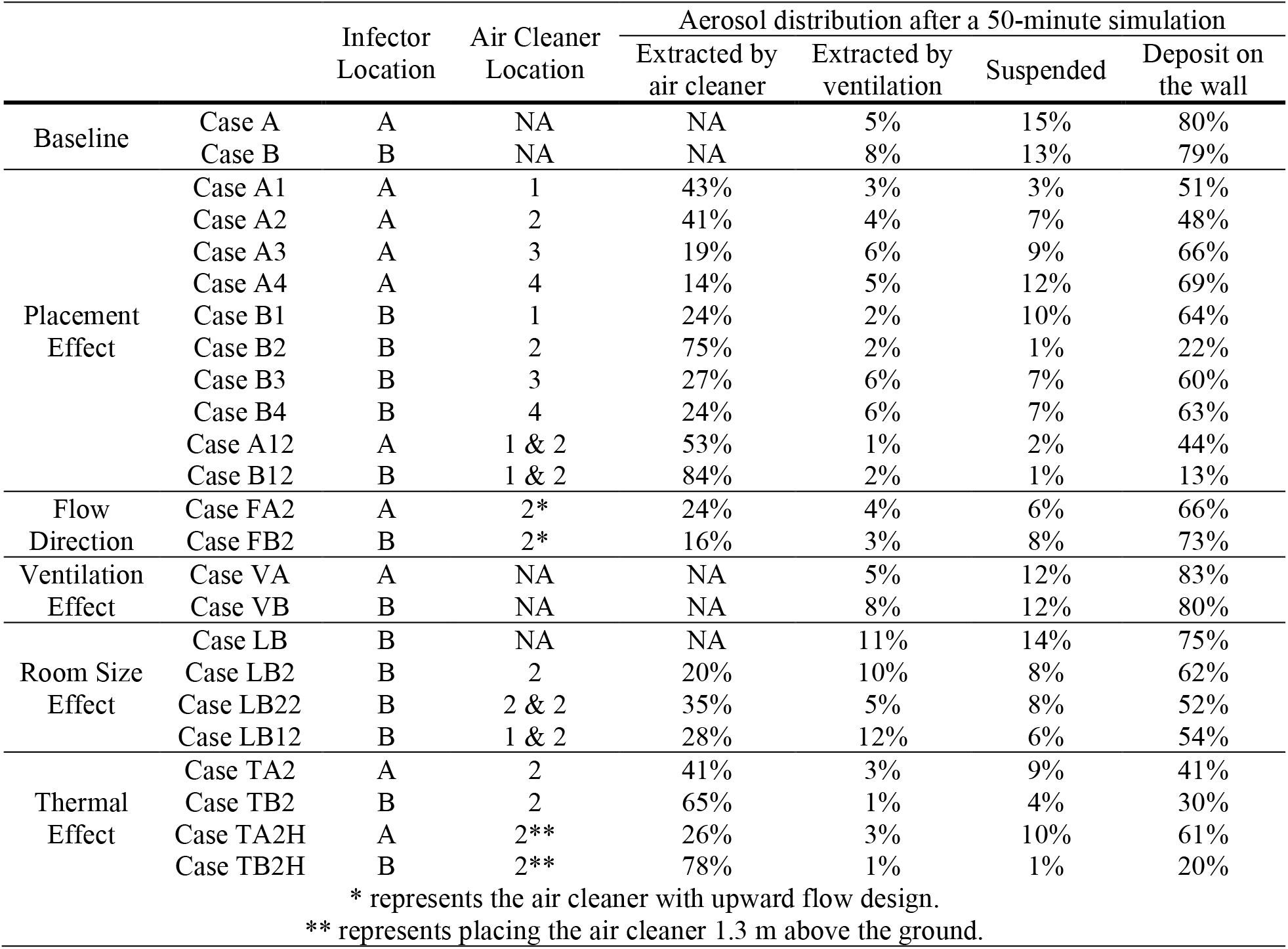
A summary of all the simulation case setups and the corresponding particle (aerosol) distribution after a 50-minute simulation period. Note that the particle distribution includes the percentages of particles that extracted by air cleaners, by the horizontal unit ventilator (HUV), suspended in the air, and deposit on the wall after 50-minute simulation.

|  |  | Infector Location | Air Cleaner Location | Aerosol distribution after a 50-minute simulation |  |  |  |
| --- | --- | --- | --- | --- | --- | --- | --- |
|  |  |  |  | Extracted by air cleaner | Extracted by ventilation | Suspended | Deposit on the wall |
| Baseline | Case A | A | NA | NA | 5% | 15% | 80% |
|  | Case B | B | NA | NA | 8% | 13% | 79% |
| Placement Effect | Case A1 | A | 1 | 43% | 3% | 3% | 51% |
|  | Case A2 | A | 2 | 41% | 4% | 7% | 48% |
|  | Case A3 | A | 3 | 19% | 6% | 9% | 66% |
|  | Case A4 | A | 4 | 14% | 5% | 12% | 69% |
|  | Case B1 | B | 1 | 24% | 2% | 10% | 64% |
|  | Case B2 | B | 2 | 75% | 2% | 1% | 22% |
|  | Case B3 | B | 3 | 27% | 6% | 7% | 60% |
|  | Case B4 | B | 4 | 24% | 6% | 7% | 63% |
|  | Case A12 | A | 1 & 2 | 53% | 1% | 2% | 44% |
|  | Case B12 | B | 1 & 2 | 84% | 2% | 1% | 13% |
| Flow Direction | Case FA2 | A | 2* | 24% | 4% | 6% | 66% |
|  | Case FB2 | B | 2* | 16% | 3% | 8% | 73% |
| Ventilation Effect | Case VA | A | NA | NA | 5% | 12% | 83% |
|  | Case VB | B | NA | NA | 8% | 12% | 80% |
| Room Size Effect | Case LB | B | NA | NA | 11% | 14% | 75% |
|  | Case LB2 | B | 2 | 20% | 10% | 8% | 62% |
|  | Case LB22 | B | 2 & 2 | 35% | 5% | 8% | 52% |
|  | Case LB12 | B | 1 & 2 | 28% | 12% | 6% | 54% |
| Thermal Effect | Case TA2 | A | 2 | 41% | 3% | 9% | 41% |
|  | Case TB2 | B | 2 | 65% | 1% | 4% | 30% |
|  | Case TA2H | A | 2** | 26% | 3% | 10% | 61% |
|  | Case TB2H | B | 2** | 78% | 1% | 1% | 20% |
| * represents the air cleaner with upward flow design. |  |  |  |  |  |  |  |
| ** represents placing the air cleaner 1.3 m above the ground. |  |  |  |  |  |  |  |

**FIG. 3.**
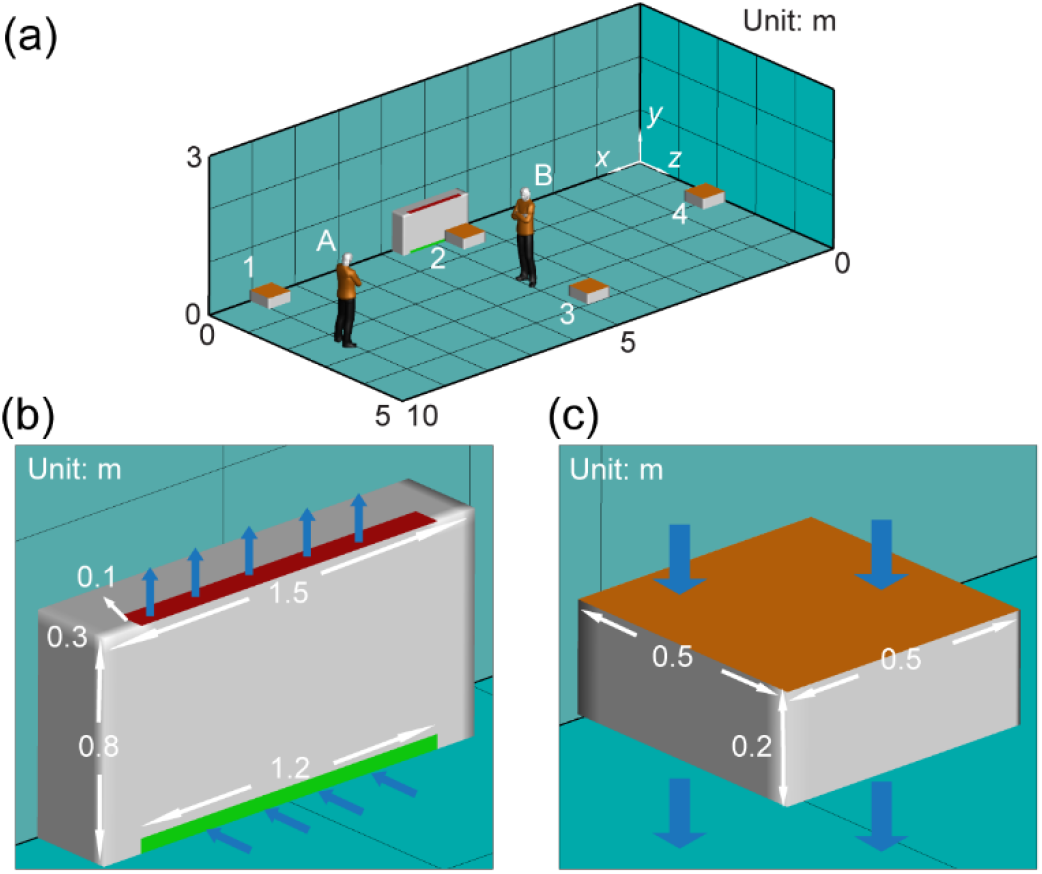
Schematics showing (a) the computational domain and locations of infectors and box fan air cleaners in the classroom, (b) the setup of horizontal unit ventilator (HUV), and (c) the model of box fan air cleaner used in the simulation.

To investigate the air cleaner placement effect, a 0.5 × 0.5 × 0.3 m^3^ cuboid located 0.3 m above the ground is used to model the box fan air cleaner in the simulation (Figure 3c). The upper surface is the inlet of the air cleaner. The profile is set according to the measurements mentioned earlier. As shown in Figure 3a, two infector locations (i.e., Locations A and B), and four air cleaner locations, i.e., in the front corner of the classroom (Location 1), in the middle of the classroom near the HUV (Location 2) and away from the HUV (Location 3), and in the back of the classroom (Location 4), in total eight cases are simulated. To study air cleaner flow direction effect, two additional cases, corresponding to two infector locations and the air cleaner placed in the middle of the classroom close to the HUV with the upward flow design (opposite to the previous cases) are included. For enhanced ventilation cases, the flow rate of HUV is increased to achieve an increase of effective air changes from 2 ACH to 5 ACH with no air cleaner added in the simulation. To further examine the room size effect, we simulate cases using a computational domain of 10 ×10 ×3 m^3,^ which doubles the size of other cases. In this simulation, the flow rate of HUV is also doubled to maintain an air exchange rate of 2 ACH for better comparison. Finally, in the cases studying the thermal effect, a 1.75 ×0.5 ×0.25 m^3^ cuboid is used to represent a simplified thermal manikin. The surface of the manikin is set to be 30 °C, the respiratory flow is set to be 34 °C^43^, while the temperature of HUV flow is 44 °C^44^. The flow rate of the respiratory flow is 2 ×10^−4^ m^3^/s based on experiment data^29^ for all the simulation cases.

To characterize the risk of encountering virus-containing particles at a given location, we use the risk index introduced by Shao et al. ^29^, denoted as *I*_risk_. It is the total number of particles passing through a given location throughout the entire duration of the simulation and can be formulated as function of spatial location *x* below:

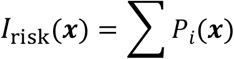

where *P*_*i*_ is defined as:

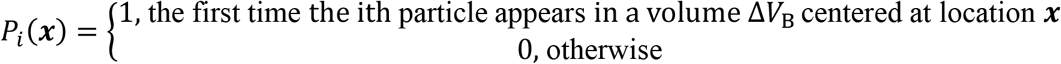

Evidently, the choice of Δ*V*_B_ influences the absolute values of *I*_risk_. Here we choose Δ*V*_B_ to be 2 × 2 × 2 c m^3^, approximating the breathing zone characterized in the schlieren imaging experiments conducted in Shao et al. ^29^ It is worth noting that the breathing zone and corresponding Δ*V*_B_ can vary substantially under different breathing conditions and across different individuals, influence the absolute values of *I*_risk_. Therefore, we mainly rely on the relative change in *I*_risk_ to evaluate the variation of airborne transmission risk under different conditions. In addition, the spatial averaged *I*_risk_ (i.e., *Ī* _risk_) along each direction (*x, y*, or *z*) is also introduced to represent the 3D distribution of *I*_risk_ in the space.

For all simulation cases, hex-core meshes generated from ICEM 18.0 are used. To determine proper mesh size for the simulation, we have conducted grid independence test for simulation Case I using two mesh sizes (1.5 million, and 2.7 million cells). Both the coarse and fine mesh yield a similar result. Therefore, we use 1.5 million cells for the remaining simulation cases related to placement effect, flow direction effect, ventilation effect, and corresponding baseline with similar settings. For simulations investigating room size effect, the total numbers of meshes are doubled for the large computational domain to maintain the mesh resolution unchanged. For the study of thermal effect, the total number of meshes are increased to 3.2 million to ensure sufficient resolution to resolve thermal plumes from the infectors.

## III. RESULTS

In this section, we will present the results of simulation cases showing the effects of placement and flow direction of box fan air cleaner on the particle removal and the corresponding distribution of airborne infection risk under the simulated classroom settings. Moreover, we will further evaluate the performance of our air cleaner for airborne risk mitigation through a comparison with the simulation cases using only enhanced ventilation (no air cleaner placed in the room). Finally, we will investigate the influence of larger room size and the inclusion of thermal effects on our simulation results. The results including the percentages of particles extracted by the air cleaner and HUV, suspended particles in the air and deposit on the surface after 50-minute simulation are summarized in Table 1.

### A. Air cleaner placement effect

The effect of the placement of the box fan air cleaner on the particle extraction and the corresponding spatial variation of airborne transmission risk in the classroom is first investigated to determine the optimal placement location under the current settings. When the infector is in the front of the classroom (Figure 4), the simulation case with no air cleaner (Figure 4a, served as the baseline) shows aerosols spread across the entire classroom, indicated by the region of *Ī* _risk_ ≥ 1 (green contour, defined as high-risk regions) extending all the way to the back of the classroom. Correspondingly, at the breathing level, the high-risk region (defined as *I*_risk_ ≥ 10, green contour) covers beyond the half of the classroom. Note that the high-risk regions here are defined in a relative sense and this definition is used consistent for all the simulations cases present in the current study. A s mentioned earlier, the absolute values of *I*_risk_ can be influenced by the definition of breathing zones and its value is only used for comparison across different cases.

**FIG. 4.**
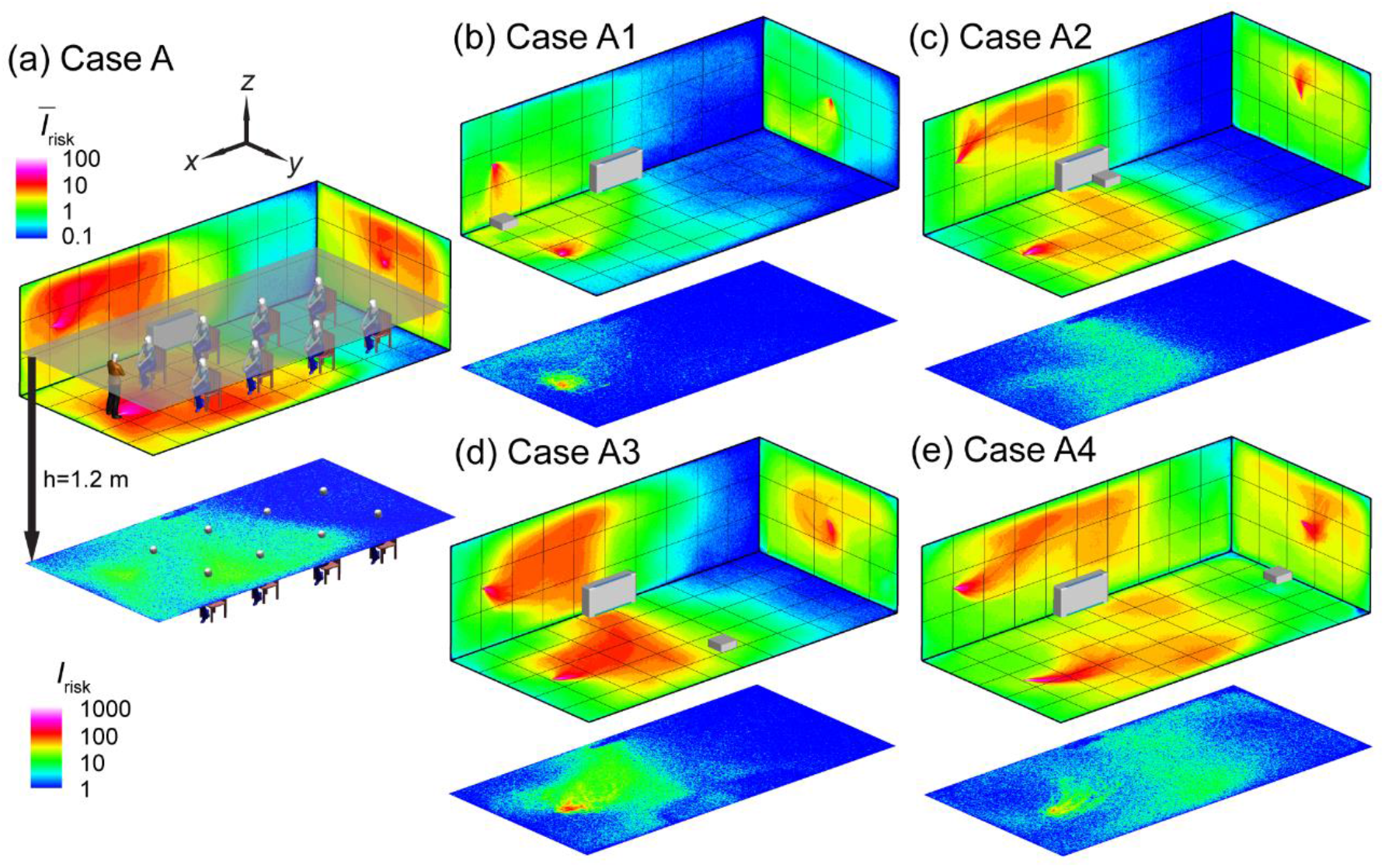
The risk index (*I*_risk_) maps of the classroom for an infector in the front of the classroom with (a) no box fan air cleaner placed (Case A), (b) the air cleaner placed in the front of the classroom (Case A1), (c) in the middle of the classroom near the horizontal unit ventilator (HUV) (case A2), (d) in the middle but away from the HUV (Case A3), and (e) in the back of the classroom (Case A4). The wall contour maps show the spatially averaged *I*_risk_ (*Ī* _risk_) along *x, y*, and *z* directions, respectively. The *I*_risk_ distribution at *x*-*y* plane at the breathing level of a sitting individual (1.2 m) are also provided. The contour of *Ī* _risk_ ≥ 1 and *I*_risk_ ≥ 10 mark the regions of high-risk (relatively) in the space. The *I*_risk_ scales are consistent between different figure types but not between the two types: spatially averaged (top) and breathing level sections (bottom).

When an air cleaner is placed near the infector (Figure 4b), the spread of aerosols is almost confined to half of the classroom (i.e., the region corresponds to *Ī* _risk_ ≥ 1). Accordingly, at the breathing level, the high-risk region is limited to an area of ∼ 1 m around the infector. In comparison, with the air cleaner moving near the HUV in the middle of the classroom (Figure 4c), although its performance in term of lowering *Ī* _risk_ and *I*_risk_ at the breathing level is reduced, but there is still considerable decrease of *Ī* _risk_ and *I*_risk_ compared with the baseline case. When the air cleaner is shifted away from the HUV in the middle (Figure 4d), the performance of the air cleaner further drops, with an enlarged area of high-risk region in both *Ī* _risk_ and *I*_risk_ maps. Finally, placing the air cleaner in the back of the classroom (Figure 4e) shows the lowest performance in suppressing *Ī* _risk_ and *I*_risk_, potentially due to the air cleaner locating farther from both the infector and HUV compared with all the other air cleaner simulation cases. Correspondingly, similar trends are observed in terms of percentages of aerosols extracted by the air cleaner and suspended aerosols among all the simulation cases with different air cleaner placements (Table 1). Specifically, when the air cleaner is placed close to the infector, it extracts 43% aerosols with only 3% suspended in the air after a 50-minute run, in comparison with the 15% suspended aerosols in the baseline case. Moving the air cleaner near the HUV maintains the same level of air cleaner extraction rate with 7% aerosols suspended. For the other two placement locations, the air cleaner extraction rate drops below 20% but the percentages of suspended aerosols are still lower than the baseline case. For all the simulation cases, a large fraction (≳ 50%) of aerosols are found to deposit on surfaces after 50 minutes.

When the infector is placed in the middle of the classroom (Figure 5), in comparison with the corresponding simulation case (Case A) in Figure 4, the baseline case shows a reduction of aerosol spread (Figure 5a) and a decrease of the percentage of suspended aerosols (i.e., from 15% in Case A to 13% in Case B) and an increase of aerosols extracted by the HUV (i.e., from 5% in Case A to 8% in Case B), potentially associated with the infector being closer to the HUV. Similarly, due to the relocation of the infector, the location where the air cleaner has the best performance is shifted (from Case A1) to the middle near the HUV (Case B2). At this location, owing to its proximity to both the infector and HUV, the air cleaner can extract 75% of aerosols and leave only 1% aerosols suspended after 50 minutes (Table 1), and correspondingly limit the high-risk regions to ∼ 1 m around the infector (Figure 5c). Remarkably, as the air cleaner is moved away from the HUV but remains in the proximity of the infector (Case B3), its performance drops significantly with the air cleaner extraction down to 25% and suspended percentage up to 7%, leading to wider spread of aerosols as shown in both *Ī* _risk_ and *I*_risk_ maps at the breathing level (Figure 5d). The performance for the air cleaner located in the front (Case B1, Figure 5b) and back (Case B4, Figure 5e) of the classroom are similar but substantially lower compared to the two previous locations. Nevertheless, the risk levels for these two cases are still considerably lower than that for the baseline case (Figure 5a), particularly in the vicinity of the infector.

**FIG. 5.**
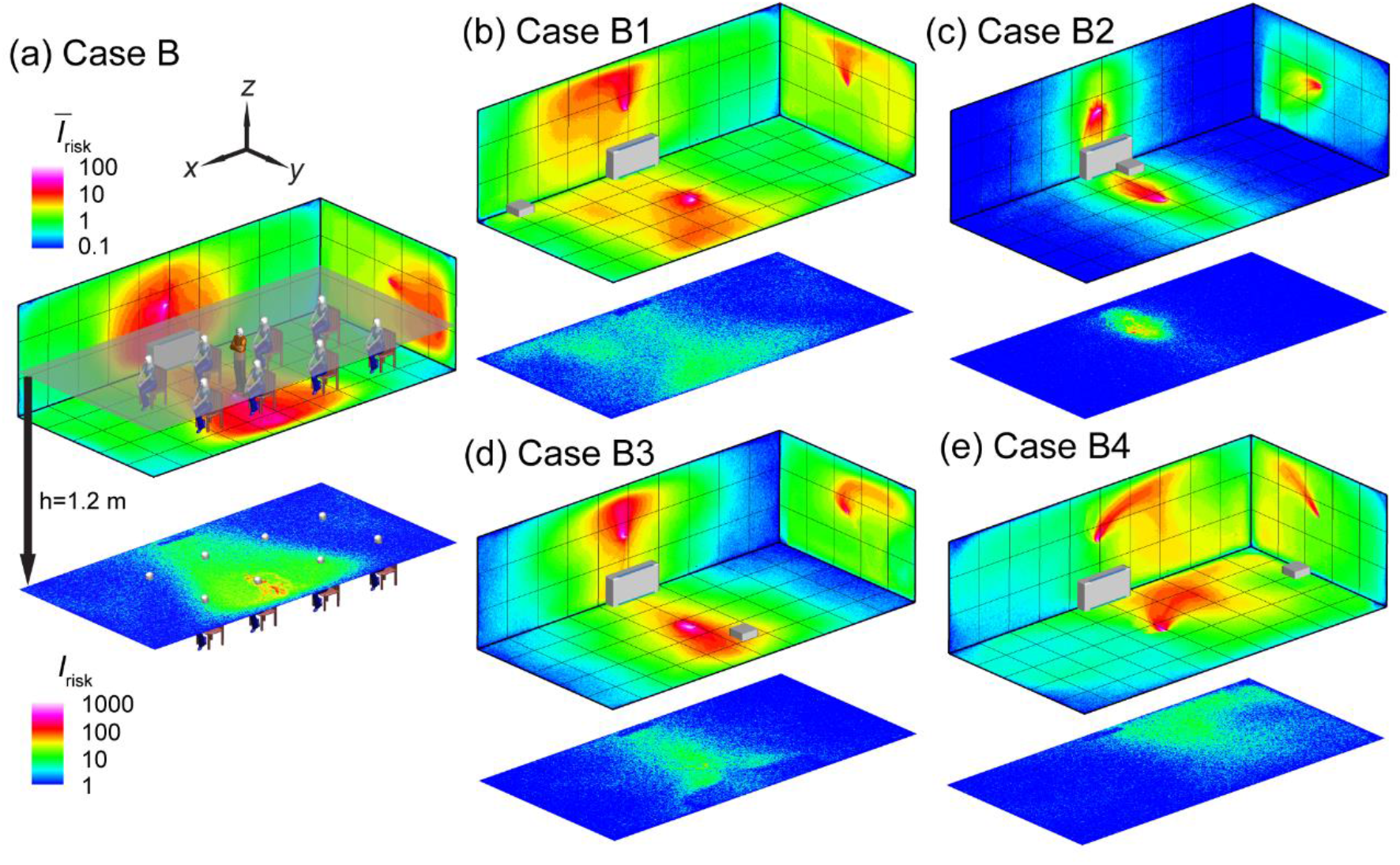
The *I*_risk_ maps of the classroom for an infector in the middle of the classroom with (a) no box fan air cleaner placed (Case B), (b) the air cleaner placed at in the front (Case B1), (c) in the middle of the classroom near the horizontal unit ventilator (HUV) (case B2), (d) in the middle but away from the HUV (Case B3), and (e) in the back of the classroom (Case B4).

Based on the abovementioned investigation on air cleaner placement effect, it can be concluded that placing the air cleaner near the infector (i.e., Case A1 and Case B2) always yields the best performance. Comparing these two cases with their corresponding baseline cases (Figure 6a, and 6d), adding air cleaners can lower the *I*_risk_ at the breathing level across the entire classroom except near the areas very near (< 1 m) the infector or the air cleaner due to the directional flow induced by the air cleaner. However, when the infector location is not known, a more common scenario in practice, placing the air cleaner near the existing HUV is optimal. Specifically, averaging the aerosol distribution for the two infector locations (Figures 4 and 5), the location close to the HUV in the middle yield the highest air cleaner extraction (58%) and lowest percentage of suspended aerosols (4%) among all four locations. Specifically, when the infector is in the front, placing the air cleaner near the HUV yields a decrease in high-risk region area across the room at the breathing level (Figure 6b).

**FIG. 6.**
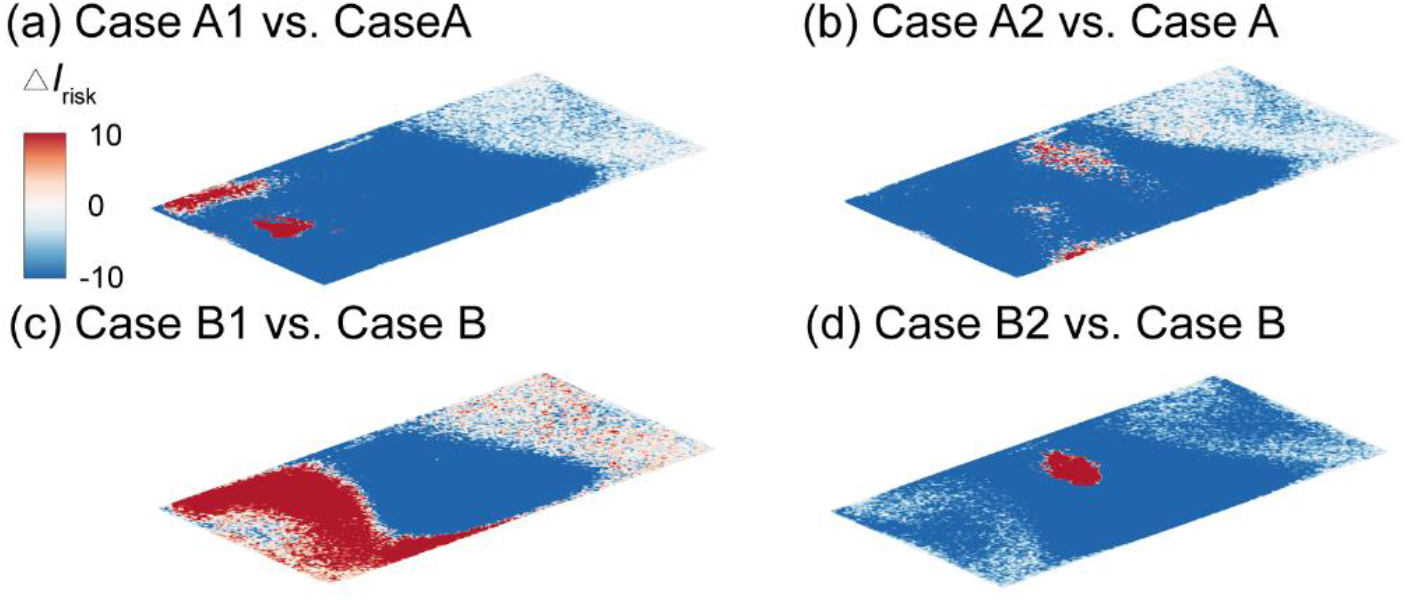
Comparison of the *I*_risk_ map (Δ*I*_risk_) at the breathing level for simulation cases with the air cleaner placed near the infector and near HUV, and the corresponding baseline case when the infector is in the front with the air cleaner placed (a) near the infector, or (b) near the HUV, and the infector is in the middle (b) with the air cleaner placed (c) near the infector, or (d) near the HUV. The Δ*I*_risk_ is defined as the *I*_risk_ of Case A1 subtracted by that of Case A for (a), Case A2 subtracted by that of Case A for (b), Case B1 subtracted by that of Case B for (c), and Case B2 subtracted by that of Case B for (d).

Furthermore, to elucidate the physical mechanism underlying the drastic performance drop of the air cleaner when its moves from near to away from the HUV (but remains in the proximity of the infector), we examine the flow field and aerosol deposition patterns for Cases B2 and B3 in comparison to Case B (Figure 7). Without an air cleaner, the streamline pattern at the *y*-*z* middle plane (across the infector and the middle of HUV) exhibits a large circulation zone away from the HUV and adjacent to right side wall (Figure 7a, highlighted by the red rectangle). Such local circulation prolongs the pathways of aerosols moving towards the HUV (illustrated by the black dashed line in Figure 7a) and hampers the extraction of aerosols by the ventilator. Instead, it increases aerosol residence time near the ceiling and right side wall, leading to a high percentage of aerosol deposition on these two walls. However, such circulation diminishes when the air cleaner is placed near the HUV (Case B2, Figure 7b, highlighted by the red rectangle). Instead, a large portion (about 70%) of the plane is dominated by downward flow towards the air cleaner and HUV (Figure 7b, highlighted by the yellow rectangle), which significantly shortens the pathway of aerosols being extracted (black dashed line in Figure 7b) and lowers their residence time near the walls. Accordingly, aerosol deposition on the ceiling and right side wall is also largely reduced. In contrast, when the air cleaner is moved away from the HUV (Case B3), the large local circulation zone re-emerges with its center shifts closer to the ceiling (Figure 7c, highlighted by the red rectangle) in comparison to that in the baseline case (Figure 7a). In addition, a small circulation zone appears in the bottom right corner, associated with the interaction between the air cleaner induced flow field and the large circulation caused by the HUV. These circulations hinder the ability of aerosols being directly transported from the infector to the air cleaner (illustrated by the long and twisted black dashed line in Figure 7c), lowering its performance drastically. These circulations also enhance the deposition of aerosols, particularly, on the right side wall near the air cleaner.

**FIG. 7.**
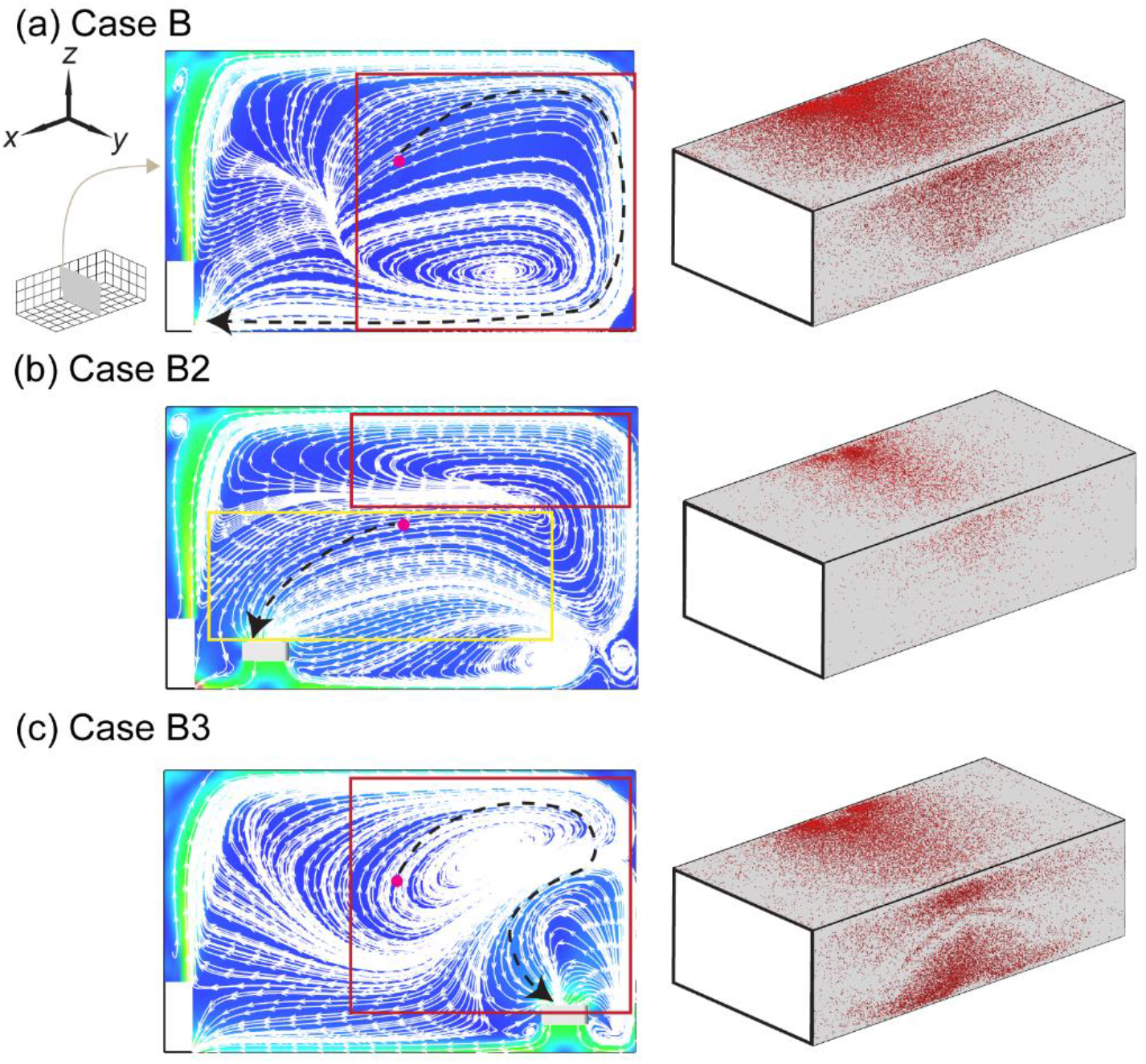
Streamline flow map at the middle *y*-*z* plane (left) and aerosol wall deposition on the ceiling and right side walls (right) for (a) Case B, (b) Case B2, and (c) Case B3. The inset figure in (a) illustrating the positions of planes shown in the figures. The magenta dot represents the inject location and the black dashed lines in the streamline maps are used to illustrate potential pathways of the aerosols being extracted by the HUV or air cleaner.

Simulations are also conducted to investigate the effectiveness of risk mitigation using multiple box fan air cleaners. Here we place one air cleaner at each of the two locations (i.e., in the front and the middle of the classroom near the HUV) that yield the best performance among all the four locations examined above and simulate for the infector in the front (Figure 8a) and the middle of the classroom (Figure 8b). For both cases, as shown in Figure 9, the increase of the number of air cleaners can lead to further reduction of high-risk regions in the entire classroom (*Ī* _risk_) and at the breathing level (*I*_risk_). Accordingly, when the infector is in the front, for the best air cleaner placement (i.e., Case A1), adding an air cleaner near the HUV can increase the percentage of aerosols extracted by air cleaners from 43% to 53% and lower the suspended aerosols from 3% to 1% (i.e., Case A1 vs Case A12). When the infector is in the middle with an air cleaner near the HUV (i.e., Case B2), the addition of an air cleaner to the front increases the air cleaner extraction from 75% to 84% and but does not lead to appreciable change in the suspended particle percentage (i.e., Case B2 vs Case B12).

**FIG. 8.**
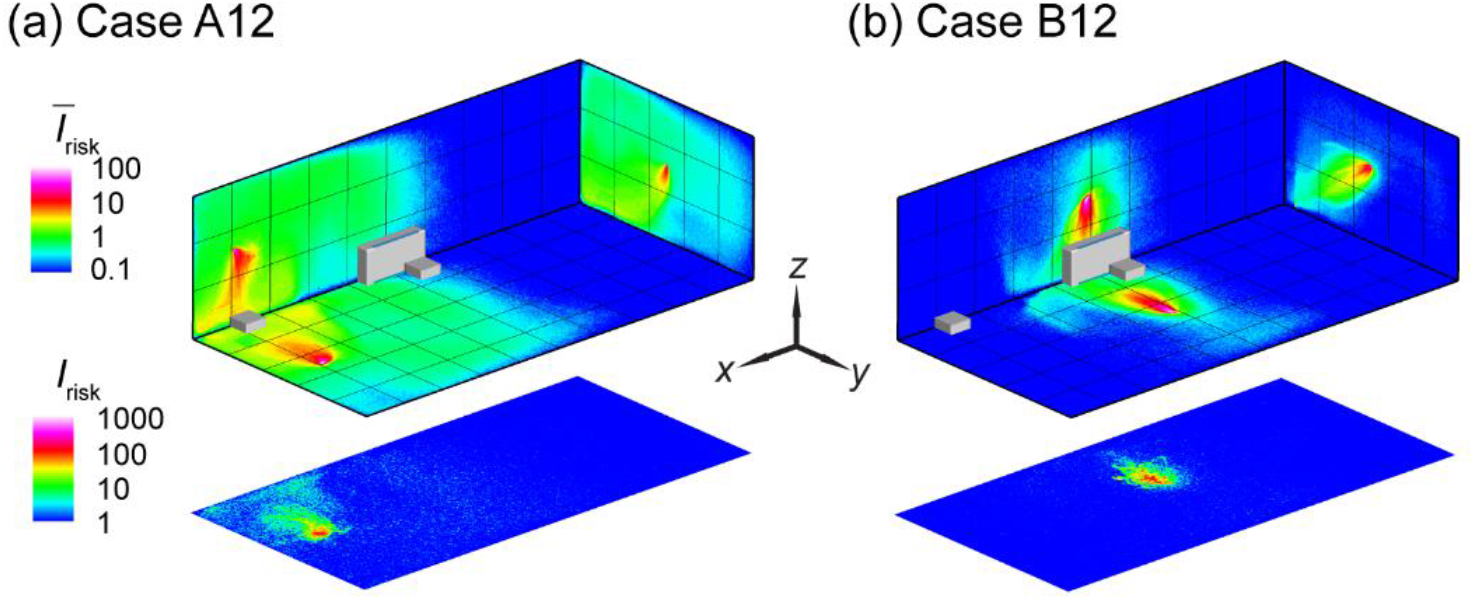
The *I*_risk_ maps of the classroom with two box fan air cleaners for an infector (a) in the front (Case A12) and (b) the middle (Case B12) of the classroom. The two air cleaners are placed in the front and the middle near the HUV of the classroom, respectively.

**FIG. 9.**
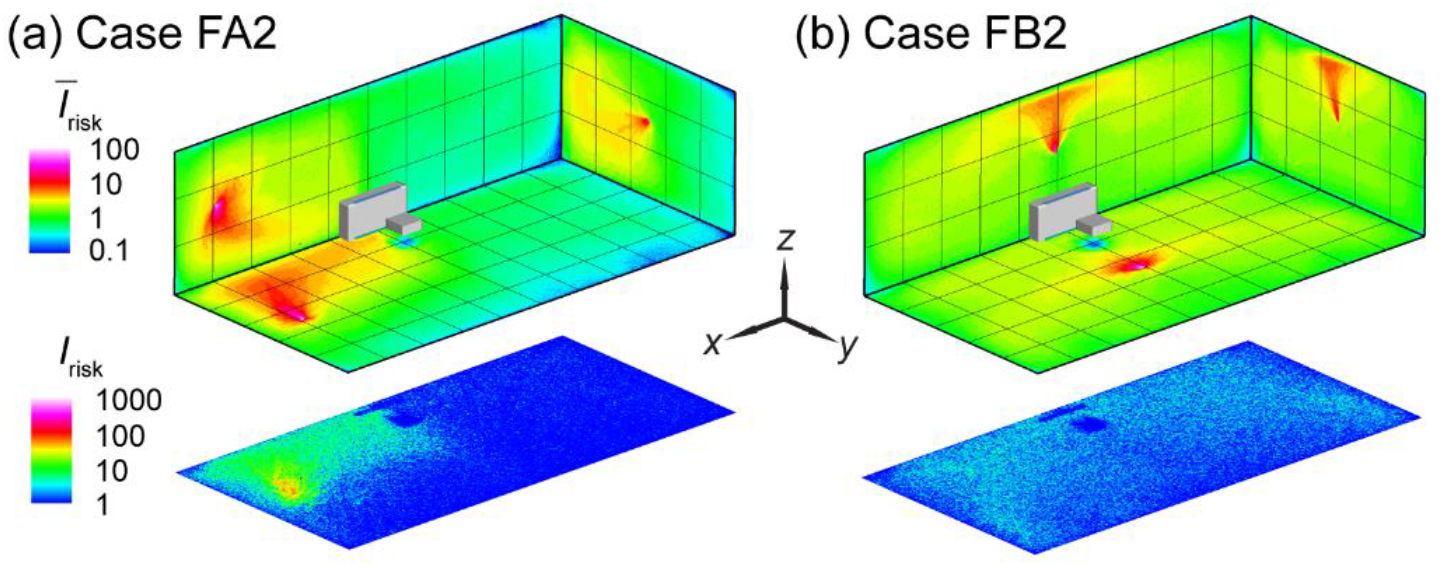
The *I*_risk_ maps of the classroom with a flipped box fan air cleaner (i.e., upward flow design) for an infector (a) in the front (Case FA2) and (b) the middle (Case FB2) of the classroom.

### B. Air cleaner flow direction effect

The design of commercial air purifiers varies substantially across different manufacturers and models. Some of them uses an “upward flow” design, such as Molekule Air, Dyson Pure Cool TP04, and Honeywell HPA600B, in which the air cleaner sucks in contaminated air at the bottom while releasing clean air on the top. Others, including Oransi ERIK650A, employs a “downward flow” to gather polluted air on the top then discharge clean air from the bottom. Comparatively, for our box fan air cleaners, all the simulation cases presented above use the downward flow design. However, to evaluate the optimal flow direction for the box fan air cleaner, additional simulations are conducted using the upward flow design with the flow inlet surface facing downward. Based on the previous findings on air cleaner placement effect, only the optimal location, i.e., the location close to the HUV, is selected for this simulation.

In comparison to the downward flow design cases (Figure 4b and Figure 5b), upward flow design generally yields a decrease in performance. In particular, when the infector is in the middle near the air cleaner, the reverse of the flow direction leads to a substantial increase in aerosol spread, evidenced from the expansion of high-risk regions to the entire classroom at the breathing level (Figure 9b). Correspondingly, the suspended aerosols percentage increases from 1% to 8% with a steep drop of aerosols extracted by the air cleaner (from 75% to 16%). Such reduction in performance is manifested from the larger portion of red areas in the Δ*I*_risk_ maps at the breathing level (Figure 10b). In comparison, the performance drop is less severe when the infector is in the front of the classroom away from the air cleaner, and the suspended aerosols stays close to the level of downward flow case. Nevertheless, a considerable decay in the aerosols extracted by the cleaner is observed (from 41% to 24%) with an elevated aerosol spread (Figure 9a).

**FIG. 10.**
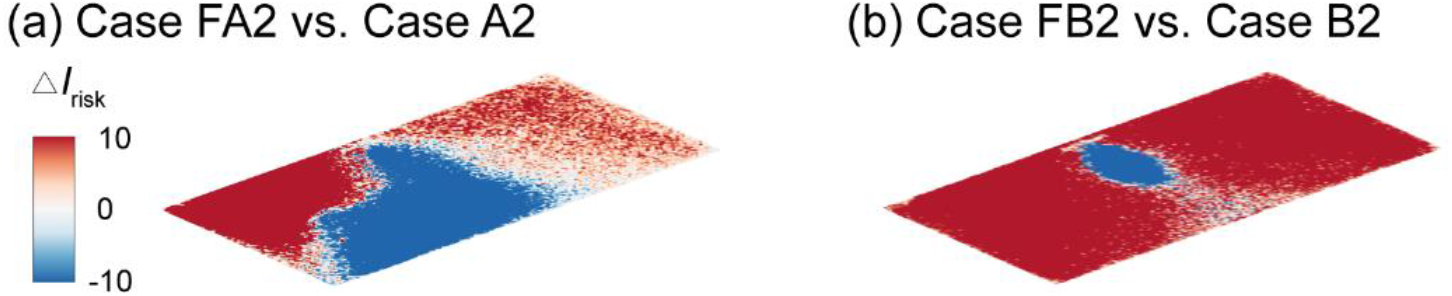
Comparison of the *I*_risk_ map (Δ*I*_risk_) at the breathing level between the downward and upward flow cases for the infector is (a) in the front and (b) in the middle of the classroom with the air cleaner located near the HUV. The Δ*I*_risk_ is defined as the *I*_risk_ of Case FA2 subtracted by that of Case A2 for (a) and the *I*_risk_ of Case FB2 subtracted by that of Case B2 for (b).

To elucidate the physical mechanism behind the significant performance reduction associated with the change of inflow direction, we investigate the flow field and aerosol deposition patterns for Case FB2 in comparison with Case B2. Specifically, when the flow design is upward, a large portion of the plane is governed by upward flow away from the HUV and the air cleaner (Figure 11, highlighted by the red rectangle) instead of dominated by the flow towards the HUV and the air cleaner in the downward design case (Figure 7b). Such flow field changes due to the flip of the flow direction significantly extends the pathway of aerosols being extracted (black dashed line in Figure 11) and increase their residence time near the walls, leading to a significant increase of the aerosol deposition (from 22% to 73%). Correspondingly, aerosol deposition on the ceiling and right side wall is increased as well (Figure 11).

**FIG. 11.**
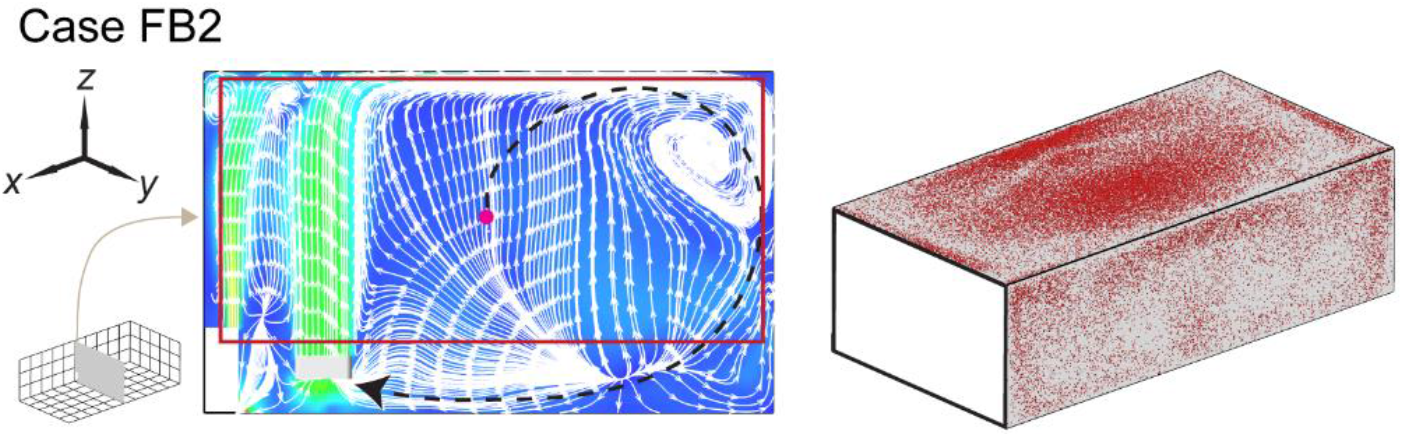
Streamline flow map at the middle *y*-*z* plane (left) and aerosol wall deposition on the ceiling and right side walls (right) for Case FB2. The magenta dot represents the inject location and the black dashed lines in the streamline maps are used to illustrate potential pathways of the aerosols being extracted by the HUV or air cleaner.

### C. Enhanced ventilation effect

A common recommendation for risk mitigation in poorly ventilated spaces is to increase ventilation rate^18^ in order to get a higher effective air changes, typically to at least 5 ACH^18^. Therefore, additional simulations are conducted to evaluate the performance of enhanced ventilation in comparison with that of placing box fan air cleaners. Here we simulate a classroom with ventilation enhanced to 5 ACH but no air cleaner for the infector in the front (Case VA, Figure 13a) and in the middle (Case VB, Figure 13b). Compared with the baseline cases at lower ventilation of 2ACH (Figure 4), ventilation enhancement can lead to reduction in the high-risk regions (Figure 12) and suspended aerosol percentage (from 15% to 8% for the infector in the front, and from 12% to 7% for the infector in the middle). However, in comparison to the optimally placed air cleaner solution (Cases A1 and B2), the performance of enhanced ventilation is significantly lower. Such discrepancy is evidenced from the larger portion of red areas in the Δ*I*_risk_ maps at the breathing level (Figure 13), which indicates an increase in risk level when enhanced ventilation case is compared with its corresponding air cleaner case. Accordingly, the air cleaner solutions yield much lower suspended aerosols (3% for Case A1, and 1% for Case B2) versus those for enhanced ventilation cases (8% for Case VA, and 7% for Case VB). Such comparison suggests that using local air cleaners placed near the infector or ventilator is a more effective approach for risk mitigation than simply enhancing the flow rate of a single ventilation unit.

**FIG. 12.**
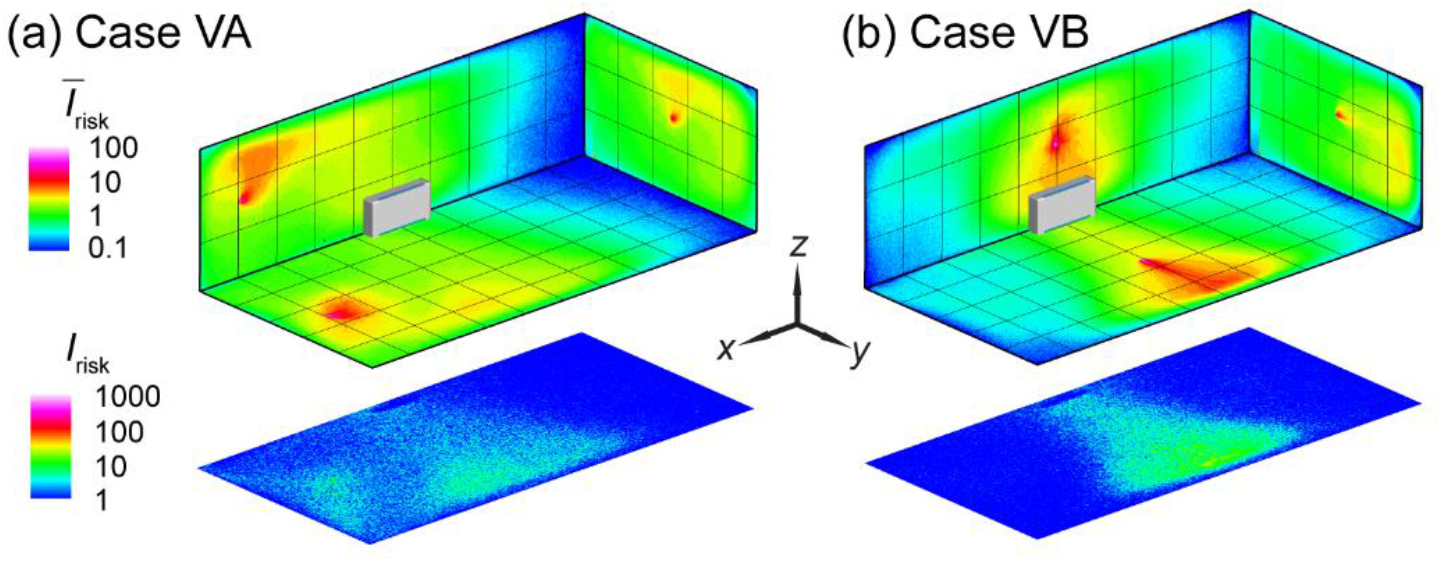
The *I*_risk_ maps of the classroom with enhanced ventilation (5 ACH) for an infector (a) in the front (Case VA) and (b) the middle (Case VB) of the classroom.

**FIG. 13.**
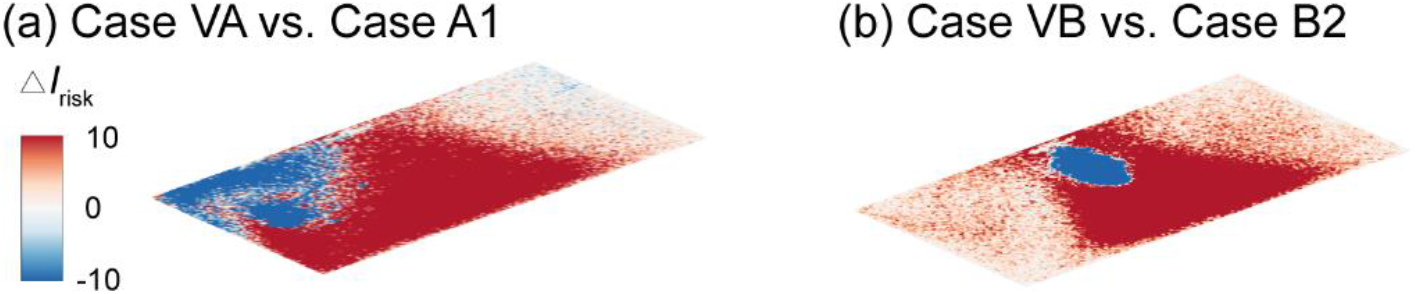
Comparison of the *I*_risk_ map (Δ*I*_risk_) at the breathing level between the enhanced ventilation and optimal placement cases for the infector (a) in the front and (b) in the middle of the classroom with the air cleaner located near the HUV. The Δ*I*_risk_ is defined as the *I*_risk_ of Case VA subtracted by that of Case A1 for (a) and the *I*_risk_ of Case VB subtracted by that of Case B2 for (b).

### D. Room size effect

In this section, we investigate the effect of classroom size on the performance of box fan air cleaners for risk mitigation since classrooms with various size are present in practice. Here we simulate a classroom with double the size of that used in previous simulations, i.e., 10 × 10 × 3 m^3^ (versus 10 × 5 × 3 m^3^ used earlier), matching one of the common classroom sizes used in the United States. In these simulations, the air cleaner is placed at its optimal location (near the HUV) for the infector in the middle. Even without changing the flow rate in a double-sized classroom, the air cleaner can still reduce the regions of high risk, particularly at the breathing level (Figure 14b), and suspended aerosol percentage (8%), in comparison to the case without air cleaners (Figure 14a and 14% suspended aerosols). However, compared with the corresponding smaller classroom case (Case B2) which only yields 1% suspended aerosols, the performance of air cleaner drops with increasing room size.

**FIG. 14.**
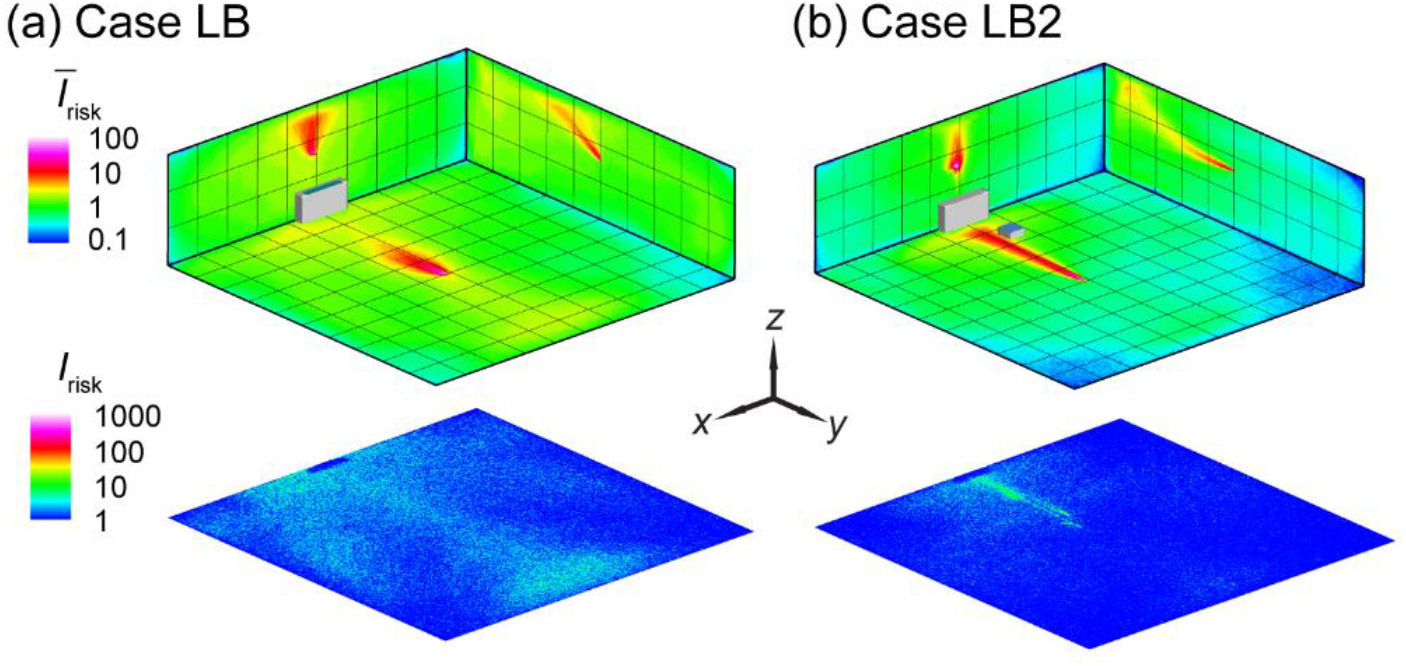
The *I*_risk_ maps of the classroom for an infector in the middle of the classroom with (a) no box fan air cleaner placed (Case LB served as a baseline) and (b) the air cleaner placed in the middle near the HUV (Case LB2).

Subsequently, to further reduce aerosol spread with increasing room size, we use simulations to examine and compare the effectiveness of two approaches, i.e., increasing air cleaner flow rate and adding more air cleaners. Specifically, two simulation cases are investigated for the infector in the middle, i.e., one that doubles the flow rate of air cleaner near the HUV and the other that adds an air cleaner in the front. Remarkably, doubling the air cleaner flow rate does not lead to appreciable reduction in suspended aerosol percentage (still 8%), but in contrary (compared with the lower rate) causes more spread of aerosols at the breathing level (Figure 15a) compared with the corresponding lower flow rate case (Figure 15b). In contrast, placing two air cleaners at lower flow rate can reduce high-risk regions at the breathing level (Figure 15b) and lower the suspended aerosols (from 8% to 6%). This result suggests that it is more effective to distribute air cleaners to multiple locations than to simply increase the flow rate of a single air cleaner or ventilation unit for risk mitigation in large size rooms.

**FIG. 15.**
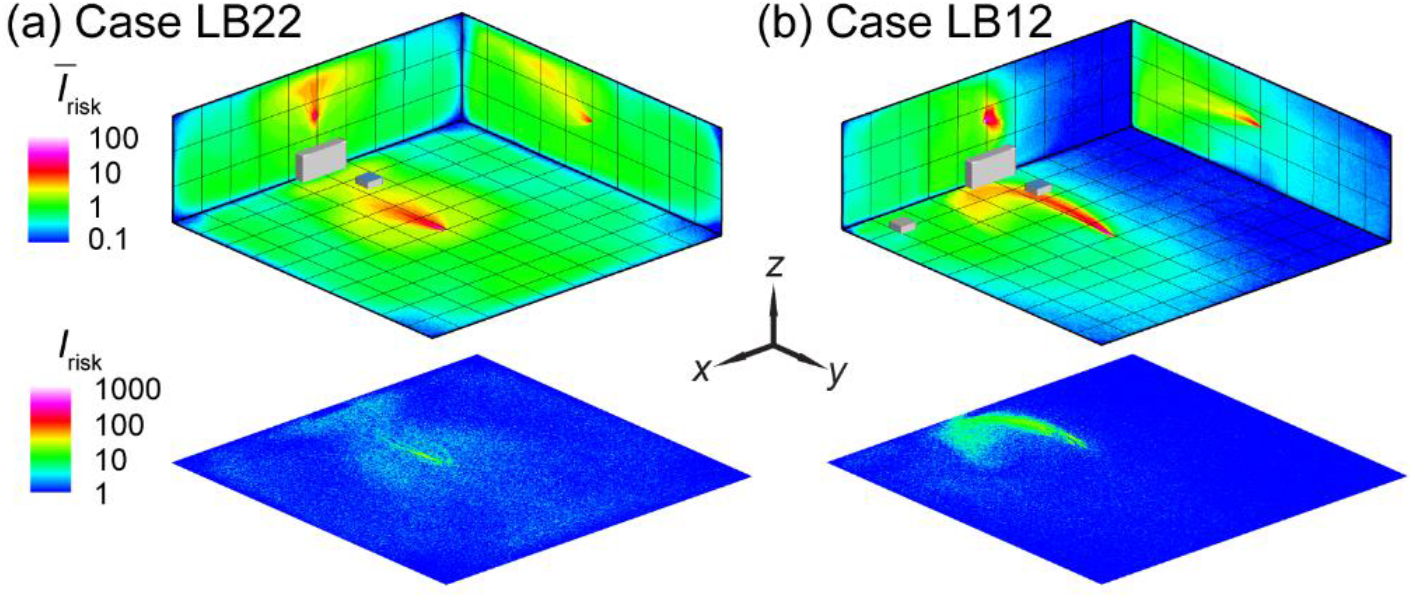
The *I*_risk_ maps of the classroom for an infector in the middle of the classroom with (a) a single air cleaner placed near the HUV with double the flow rate of the previous simulation cases (Case LB22), and (b) one air cleaner near the HUV and the other in the front (Case LB12).

### E. Thermal effect

As shown in the literature, the temperature difference among ventilation air, human surface temperature, and ambient room air can influence the spread of aerosols in indoor spaces. Particularly, the heated classrooms in winter and air-conditioned classrooms in summer may yield considerable temperature gradient which could lead to a thermal flow that is comparable to or more dominant than ventilation flow in, especially, poorly ventilated spaces. Therefore, we conduct the simulation under a simplified scenario representing a heated classroom in winter. When the thermal effect associated with human thermal plume and hot ventilation is included in the simulation, the performance of box fan air cleaners drops, manifested as an increase in high-risk regions in the *Ī* _risk_ and *I*_risk_ maps (Figure 16a *vs* Figure 4c and Figure 16b *vs* Figure 5c for the infector at in the front and in the middle of the classroom, respectively). Such increase is illustrated more clearly in the Δ *I*_risk_ map at the breathing level, corresponding to the larger area of red contours in comparison to that of blue in Figure 17. Correspondingly, with the inclusion of thermal effect, the suspended aerosol percentage increases from 7% to 9 % and from 1% to 4% for the infector in the front and the middle, respectively. It is worth noting that the decrease in air cleaner performance is more substantial when the air cleaner is located farther away from the infector. We attribute such decrease to the change in flow patterns associated with thermal effect. Specifically, the flow induced by the thermal gradient causes the formation of large recirculation adjacent to the ceiling (Figure 18, highlighted by the red rectangle). The aerosols produced by the infector tend to move upwards due to the thermal plume and be trapped in this circulation, thus have higher chance to deposit on the wall and disperse instead of directly transport towards the air cleaner (Figure 18).

**FIG. 16.**
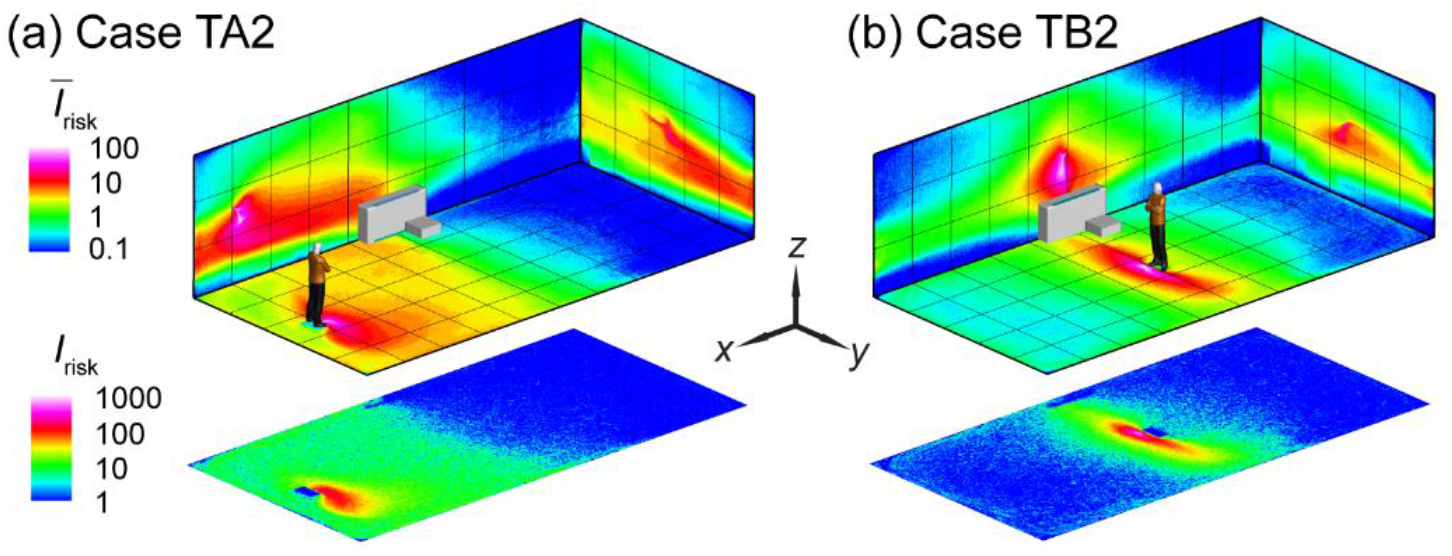
The *I*_risk_ maps of the classroom when the thermal effect associated with human thermal plume and hot ventilation is considered for the infector (a) in the front (Case TA2) and (b) the middle of the classroom with the air cleaner near the HUV (Case TB2).

**FIG. 17.**
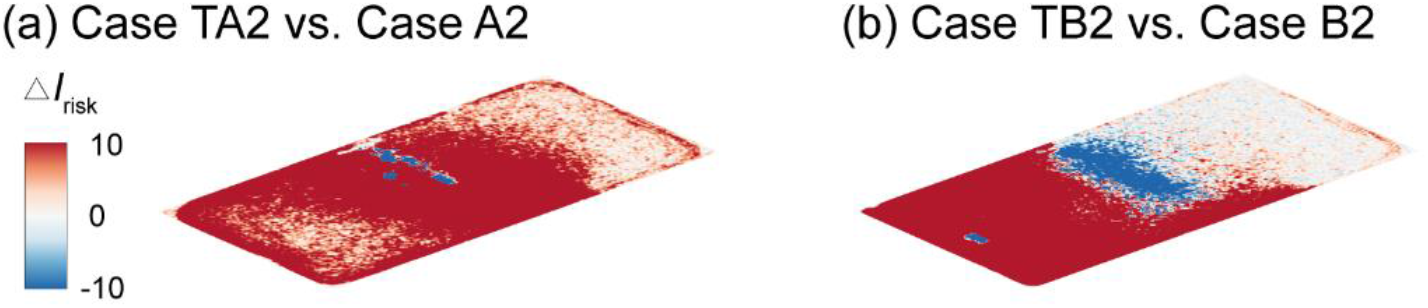
Comparison of the *I*_risk_ map at the breathing level between the simulation cases with and without consideration of thermal effect for the infector (a) in the front and (b) in the middle of the classroom with the air cleaner near the HUV. The Δ*I*_risk_ is defined as the *I*_risk_ of Case TA2 subtracted by that of Case A2 for (a) and the *I*_risk_ of Case TB2 subtracted by that of Case B2 for (b).

**FIG. 18.**
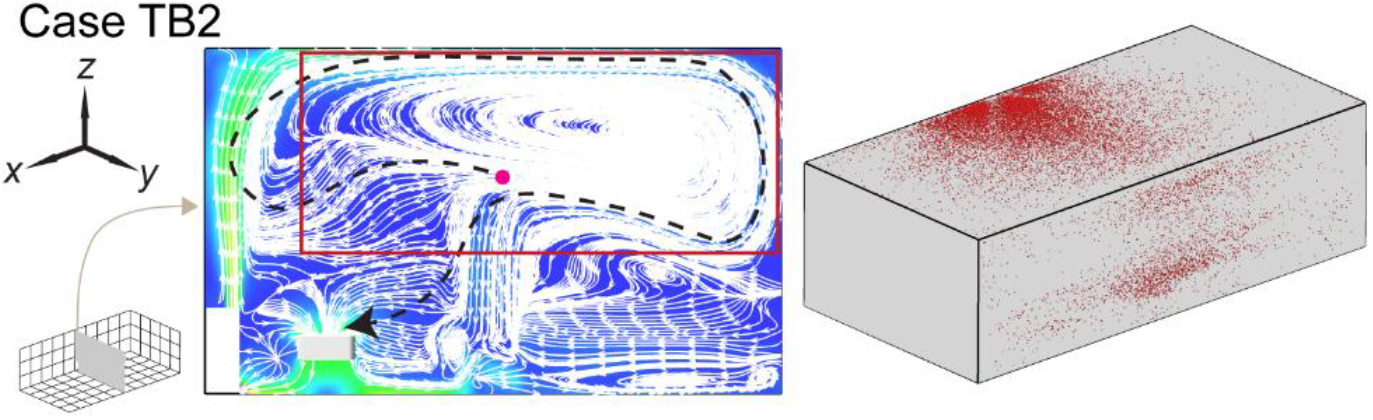
Streamline flow map at the middle *y*-*z* plane (left) and aerosol wall deposition on the ceiling and right side walls (right) for Case TB2. The magenta dot represents the inject location and the black dashed lines in the streamline maps are used to illustrate potential pathways of the aerosols being extracted by the HUV or air cleaner.

To explore whether the placement of air cleaners can be adjusted to achieve better performance under the influence of thermal gradient, we simulate additional cases in which the air cleaner is raised 1 m vertically from its original position, i.e., 1.3 m above the floor (Figure 19). As shown in Figures 19b and 20b, a clear improvement in air cleaner performance is observed for the case with the infector in the middle and the air cleaner located in proximity of the infector. This improvement is because the elevating air cleaner can take advantage of human thermal plume to improve its particle extraction (changing from 65% to 78%) and decrease the spread of aerosol transmission (Figure 20b). However, when the infector is located farther away from the air cleaner (Figures 19a), the performance of the air cleaner drops with elevated placement (indicated by the larger area of red contour than that of blue in Figure 20a). Such discrepancy in the performance of the elevated air cleaner between Case TA2H and TB2H is due to the fact that human thermal plume is only dominant in the vicinity of the infector and the elevated air cleaner located far away from the infector can no longer benefit from the aerosol transport by thermal updraft.

**FIG. 19.**
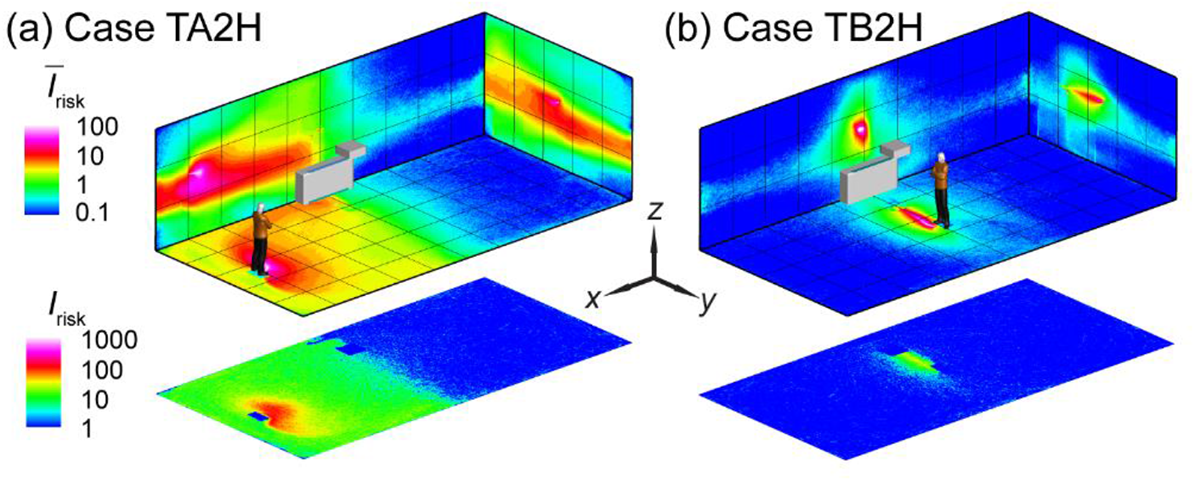
The *I*_risk_ maps of the classroom when the thermal effect is considered with the air cleaner placed near the HUV at a higher elevation (1.3 m above the floor) compared with previous simulation cases (0.3 m above the floor) for an infector (a) in the front (Case TA2H) and (b) in the middle (Case TB2H) of the classroom.

**FIG. 20.**
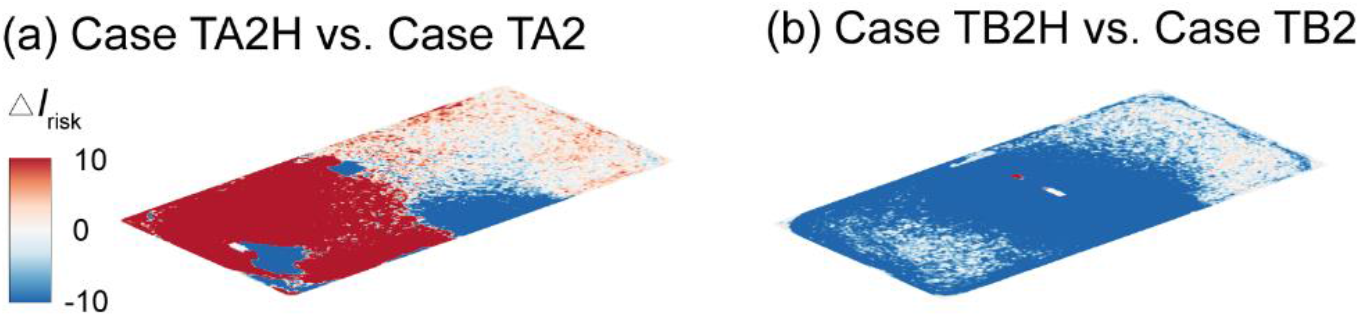
Comparison of the *I*_risk_ map at the breathing level between the simulation cases with an air cleaner placed 1.3 m and 0.3 m above the floor near the HUV for the infector (a) in the front and (b) in the middle of the classroom. The Δ*I*_risk_ is defined as the *I*_risk_ of Case TA2H subtracted by that of Case TA2 for (a) and the *I*_risk_ of Case TB2H subtracted by that of Case TB2 for (b).

## IV. Conclusion & Discussion

Using computational fluid dynamics, we provide a systematic investigation of airborne transmission in a poorly ventilated classroom and evaluate the performance of low-cost box fan air cleaners for risk mitigation. The classroom is modeled with a single horizontal unit ventilator (HUV) operating at an air exchange rate of ∼2 ACH, representing the ventilation setting in a typical classroom built before 1989 ^30^. Our study shows that placing box fan air cleaners in the classroom result in a substantial reduction of airborne transmission risk across the entire space. The performance of the cleaner, in terms of its efficiency to extract aerosols and lower the percentage of suspended aerosols from potential infectors, is strongly influenced by its placement. We find that the cleaner can achieve best performance when placed near the infector. However, without knowing the location of the patient, the performance of the cleaner is optimal near the HUV. Specifically, at the optimal placement the cleaner can extract majority of aerosols emitted continuously from an asymptomatic instructor (infector) and reduce the suspended aerosols down to 1% after a 50-minute simulation of a lecture, significantly lower than the condition without the cleaner (13%). In addition, the simulations show that the air cleaner with downward flow design (i.e., the flow inlet of the cleaner facing upward) performs better than the upward flow one, resulting in a more confined high-risk region and lower percentage of suspended aerosols when situated near the infector particularly.

In comparison to raising the air exchange rate of HUV (i.e., from 2 ACH to 5 ACH), the air cleaner solution can result in higher reduction in aerosol concentration and spread in the classroom. As the classroom size increases, with placement of additional cleaners separately in the domain, air cleaner solution can still lead to a confined dispersion of aerosols and a significant reduction of the suspended aerosols. In contrast, doubling the cleaner flow rate to accommodate increasing room size may cause even more spread of aerosols at the breathing level compared with the cleaner operating at lower flow rate. When considered the thermal gradient associated with human thermal plume and hot ventilation air during cold seasons, overall performance of air cleaners drops but their efficacy in reducing aerosol spread and regions of high-risk airborne transmission still holds compared with the baseline case with only ventilation. We also find that elevating the cleaner when it is placed near an infector can increase its performance by taking advantage of human thermal plumes that drive particles moving upward.

Our work has demonstrated the various effects of implementing air cleaners in a poorly ventilated classroom that relies on a horizontal unit ventilator (HUV). The methodology and the results from our study are generally applicable for evaluating of efficacy of the air cleaner solution for other poorly ventilated spaces including aged offices, prisons, homeless shelters, etc. One of the insights of the CFD analysis is that the air cleaner can not only reduce the overall concentration of aerosols in the space, but also limit the spread. These results could conceivably be applied to other types of portable air filtration systems. A novel aspect of this study, compared to previous portable air filtration system studies, is that it does not look at a “well mixed”, more uniform distribution across the space. Instead, it explores the implications of placing portable air cleaners in a representative space as it is likely done in practice.

According to our results, placing the air cleaner next to the infector is the most effective. If the individuals (if they are asymptomatic infectors) who can impose the highest risk to others can be identified in a space, air cleaners should be placed in the proximity of these individuals to limit the spread of their emitted aerosols. In practical settings (e.g., classroom, concert, etc.), it is better to take the precaution to place air cleaners near unmasked teachers, singers, and trumpet players who can produce large number of aerosols during their activities or a new person entering a relatively quarantined group. However, when such high risk individuals and their locations cannot be identified beforehand, the best practice is to place the air cleaner near the existing ventilation system. Under such placement, air cleaner is acting as a high specification filter for the unit ventilator. It is shown from the simulation that the convection flow is enhanced when placing the air cleaner close to the existing ventilation system, thus minimizing the recirculation zone in the space, and allowing more aerosols to be entrained in the main circulation path and removed by the air cleaner. Unit ventilators are performing the job of conditioning the air and providing fresh oxygen to the room. However, many of them are not designed to take a high specification filter. By implementing the air cleaner near the existing ventilation system, the air available to the unit ventilator is filtered as to add this capability to the system. This allows the unit ventilator to continue conditioning and mixing the air without losing performance and disrupting the circulation of the room.

Two air cleaners in the room can capture a large portion of the emitted aerosols from the infector directly into the air cleaners. There is a localized high-risk region close to the infector but the rest of the space has low risks. Our results suggest that multiple air cleaners could be used to locally target and remove aerosols and limit their spread across the room. Although such deployment depends on the available resources and the type of HVAC system that is being utilized, it is a definite advantage of the low-cost box fan air cleaner as it could allow multiple deployments in a space for the same or fraction of the price as one expensive commercial air purifier when properly weighted against other factors such as noise.

It is worth noting that all the cases are simulated with an aerosol emission rate corresponding to unmasked individuals. Considering wearing mask in closed spaces such as classrooms is a suggested method which can potentially lower the aerosol emission, it is conceivable that the risk levels under masked conditions are substantially lower than those in our simulated cases. Nevertheless, we expect the spatial distribution of risk regions reported in our study will not be largely influenced by the presence of masks since the mask only affect the flow field very near the infector and the transport of aerosols in the space is dominated by the flows generated from the ventilation and cleaners. In addition, it is worth noting that wearing mask can hampers voice directivity and speech intelligbility^45^, imposing a detrimental impact on learning, particularly in large classrooms or for hearing impaired learners. As an alternative solution, placing the air cleaner close to the instructor can substantially mitigate the transmission risk without compromising on teach quality.

There are many variables that affect specific details of airflow, ventilation, and aerosol dynamics in a particular space. Our results only provide general trends and should not be treated as absolute criteria for a specific environment. For example, the particle-wall interaction model relies on commonly used assumptions^46^. Its validity on aerosol size particles relevant to disease transmission has not been fully examined, which may impose an uncertainty on the aerosol percentage present in our study. Nevertheless, the comparison in the relative percent reduction between different cases are valuable information. The thermal effects are examined in a simplified classroom setting which demonstrates that the addition of thermal plumes does not change the effectiveness of the system. However, our simulation uses a simplistic environment with uniform wall conditions and very few loads in the space (e.g., people, equipment). These simplifications may cause some difference in the flow patterns (e.g., the upper circulation zone) between our simulation and real settings. Nevertheless, the comparison of our results (thermal cases) with nonthermal cases indicates that the efficacy of our cleaners for risk mitigation remains reasonable robust against the change of flow patterns associated with thermal effect.

Correlating the modeled aerosol concentration and distribution over time to field and laboratory measurements will be important to validate the parameters and boundary conditions used in this study. In addition, a follow-up study involving a systematic comparison between experiments and CFD can help provide a deeper understanding of how well CFD modeling tools can reliably assess concentration and risk, especially for unique boundary conditions, complex thermal effects, aerosol counts, and use cases not directly addressed in this study.

## Data Availability

The data that support the findings of this study are available from the corresponding author upon reasonable request.

## ACKNOWLEDGMENTS

We acknowledge the support from Ford Motor Company for this research. We would like to thank Dr. Siyao Shao and Mr. Rafael Grazzini Placucci for conducting the flow visualization experiment. In addition, we also would like to acknowledge the support from Minnesota Supercomputing Institute (MSI) that provides computational resources.

## Notes

### Competing Interest Statement

The authors have declared no competing interest.

### Funding Statement

Ford Motor Company provides support in the process.

### Author Declarations

This research doesn't involve human being or living animals.

## References

1. L. Morawska and J. Cao, “Airborne transmission of SARS-CoV-2: The world should face the reality” Environment International 139, 105730, (2020).

2. J.A. Lednicky, M. Lauzard, Z.H. Fan, A. Jutla, T.B. Tilly, M. Gangwar, M. Usmani, S.N. Shankar, K. Mohamed, A. Eiguren-Fernandez, C.J. Stephenson, M.M. Alam, M.A. Elbadry, J.C. Loeb, K. Subramaniam, T.B. Waltzek, K. Cherabuddi, J.G. Morris, and C.Y. Wu, “Viable SARS-CoV-2 in the air of a hospital roo m with COVID-19 patients” International Journal of Infectious Diseases 100, 476–482 (2020).

3. J. Lu, J. Gu, K. Li, C. Xu, W. Su, Z. Lai, D. Zhou, C. Yu, B. Xu, and Z. Yang, “COVID-19 Outbreak Associated with Air Conditioning in Restaurant, Guangzhou, China, 2020” Emerging Infectious Diseases 26.7, 1628 (2020).

4. J.L. Santarpia, D.N. Rivera, V.L. Herrera, M.J. Morwitzer, H.M. Creager, G.W. Santarpia, K.K. Crown, D.M. Brett-Major, E.R. Schnaubelt, M.J. Broadhurst, J. v. Lawler, S.P. Reid, and J.J. Lowe, “Aerosol and surface contamination of SARS-CoV-2 observed in quarantine and isolation care” Scientific Reports 10, 1–8 (2020).

5. S.Y. Park, Y.M. Kim, S. Yi, S. Lee, B.J. Na, C.B. Kim, J. il Kim, H.S. Kim, Y.B. Kim, Y. Park, I.S. Huh, H.K. Kim, H.J. Yoon, H. Jang, K. Kim, Y. Chang, I. Kim, H. Lee, J. Gwack, S.S. Kim, M. Kim, S. Kweon, Y.J. Choe, O. Park, Y.J. Park, and E.K. Jeong, “Coronavirus disease outbreak in call center, South Korea” Emerging Infectious Diseases 26.8, 1666 (2020).

6. S.L. Miller, W.W. Nazaroff, J.L. Jimenez, A. Boerstra, G. Buonanno, S.J. Dancer, J. Kurnitski, L.C. Marr, L. Morawska, and C. Noakes, “Transmission of SARS-CoV-2 by inhalation of respiratory aerosol in the Skagit Valley Chorale superspreading event “ Indoor Air (2020).

7. Y. Shen, C. Li, H. Dong, Z. Wang, L. Martinez, Z. Sun, A. Handel, Z. Chen, E. Chen, M.H. Ebell, F. Wang, B. Yi, H. Wang, X. Wang, A. Wang, B. Chen, Y. Qi, L. Liang, Y. Li, F. Ling, J. Chen, and G. Xu, “Community Outbreak Investigation of SARS-CoV-2 Transmission among Bus Riders in Eastern China “ JAMA Internal Medicine 180.12, 1665–1671 (2020).

8. ASHRAE, “Pandemic COVID-19 and Airborne Transmission” (2020).

9. L. Morawska, J.W. Tang, W. Bahnfleth, P.M. Bluyssen, A. Boerstra, G. Buonanno, J. Cao, S. Dancer, A. Floto, F. Franchimon, C. Haworth, J. Hogeling, C. Isaxon, J.L. Jimenez, J. Kurnitski, Y. Li, M. Loomans, G. Marks, L.C. Marr, L. Mazzarella, A.K. Melikov, S. Miller, D.K. Milton, W. Nazaroff, P. v. Nielsen, C. Noakes, J. Peccia, X. Querol, C. Sekhar, O. Seppänen, S. ichi Tanabe, R. Tellier, K.W. Tham, P. Wargocki, A. Wierzbicka, and M. Yao, “How can airborne transmission of COVID-19 indoors be minimized?” Environment International 142, 105832 (2020).

10. A.K. Melikov, “Advanced air distribution: Improving health and comfort while reducing energy use” Indoor Air 26.1, 112–124 (2016).

11. Nowicki Jacqueline M. “K-12 Education: School Districts Frequently Identified Multiple Building Systems Needing Updates or Replacement. Report to Congressional Addressees. GAO-20-494” US Government Accountability Office (2020).

12. W.R. Chan, X. Li, B.C. Singer, T. Pistochini, D. Vernon, S. Outcault, A. Sanguinetti, and M. Modera, “Ventilation rates in California classrooms : Why many recent HVAC retrofits are not delivering sufficient ventilation “ Building and Environment 167, 106426 (2020).

13. J. Adelman, “Fresh air will be a hot amenity in reopened offices if companies can afford it” The Philadelphia Inquirer (2020).

14. H. Qian, Y. Li, W.H. Seto, P. Ching, W.H. Ching, and H.Q. Sun, “Natural ventilation for reducing airborne infection in hospitals.” Building and Environment 45.3, 559–565 (2010).

15. “Ventilation in Buildings” Centers for Disease Control and Prevention (CDC), (2021) https://www.cdc.gov/coronavirus/2019-ncov/community/ventilation.html

16. R.J. Shaughnessy and R.G. Sextro, “What is an effective portable air cleaning device? A review” Journal of Occupational and Environmental Hygiene 3.4, 169–181 (2006).

17. J. Curtius, M. Granzin, and J. Schrod, “Testing mobile air purifiers in a school classroom: Reducing the airborne transmission risk for SARS-CoV-2” Aerosol Science and Technology 1–8 (2021).

18. J. Allen, J. Spengler, E. Jones, and J. Cedeno-Laurent, “5-step guide to checking ventilation rates in classrooms “ Harvard Healthy Buildings program (2020). https://schools.forhealth.org/wp-content/uploads/sites/19/2021/01/Harvard-Healthy-Buildings-program-How-to-assess-classroom-ventilation-10-30-2020-EN_R1.8.pdf

19. S. Kirkman, J. Zhai, and S.L. Miller, “Effectiveness of Air Cleaners for Removal of Virus-Containing Respiratory Droplets: Recommendations for Air Cleaner Selection for Campus Spaces” (2020). https://shellym80304.files.wordpress.com/2020/06/air-cleaner-report.pdf

20. A. Tseng, “DIY air filters can be safe, simple and inexpensive. Here’s how to make one.” LA Times (2020). https://www.latimes.com/environment/story/2020-09-17/best-air-filters-sold-out-how-to-make-diy-purifier

21. H. Liu, S. He, L. Shen, and J. Hong, “Simulation-based study of COVID-19 outbreak associated with air-conditioning in a restaurant” Physics of Fluids 33.2, 023301 (2021).

22. M.-R. Pendar and J.C. Páscoa, “Numerical modeling of the distribution of virus carrying saliva droplets during sneeze and cough” Physics of Fluids 32.8, 083305 (2020).

23. Z. Li, H. Wang, X. Zhang, T. Wu, and X. Yang, “Effects of space sizes on the dispersion of cough-generated droplets from a walking person” Physics of Fluids 32.12, 121705 (2020).

24. J. Xi, X.A. Si, and R. Nagarajan, “Effects of mask-wearing on the inhalability and deposition of airborne SARS-CoV-2 aerosols in human upper airway.” Physics of Fluids 32.12, 123312 (2020).

25. A. Foster and M. Kinzel, “Estimating COVID-19 exposure in a classroom setting: A comparison between mathematical and numerical models “ Physics of Fluids 33.2, 021904 (2021).

26. Z. Lin, J. Wang, T. Yao, T.T. Chow, and K.F. Fong, “Numerical comparison of dispersion of human exhaled droplets under different ventilation methods” World Review of Science, Technology and Sustainable Development 10, 142–161 (2013).

27. Y. Zhang, G. Feng, K. Huang, and G. Cao, “Numerical Simulation of Aerosol Particles Distribution in a Classroom “ Proceedings of the 8th International Symposium on Heating, Ventilation and Air Conditioningp203–210 (2014).

28. M. Abuhegazy, K. Talaat, O. Anderoglu, S. v. Poroseva, and K. Talaat, “Numerical investigation of aerosol transport in a classroom with relevance to COVID-19” Physics of Fluids 32.10, 103311 (2020).

29. S. Shao, D. Zhou, R. He, J. Li, S. Zou, K. Mallery, S. Kumar, S. Yang, and J. Hong, “Risk assessment of airborne transmission of COVID-19 by asymptomatic individuals under different practical settings “ Journal of Aerosol Science 151, 105661 (2021).

30. “School HVAC Design Manual” DAIKIN, (2014)

31. A. Abraham, R. He, S. Shao, S. Santosh Kumar, C. Wang, B. Guo, M. Trifonov, R. Grazzini Placucci, M. Willis, and J. Hong, “Risk Assessment and Mitigation of Airborne Disease Transmission in Orchestral Wind Instrument Performance” (2020). https://doi.org/10.1101/2020.12.23.20248652

32. S.R. Narayanan and S. Yang, “Airborne transmission of virus-laden aerosols inside a music classroom: Effects of portable purifiers and aerosol injection rates “ Physics of Fluids 33, 033307 (2021)

33. “How to Assemble a COVID-19 Air Filtration Box Fan “ https://advocate.socialchorus.com/ford/blueovalnow/articles/how-to-assemble-a-covid-19-air-filtration-box-fan?5d08ebff=fa2a1297.

34. “Method of Testing General Ventilation Air-Cleaning Devices for Removal Efficiency by Particle Size” ASHRAE Standing Standard Project, (2017).

35. “ANSI/AHAM AC-1-2019” Association of Home Appliance Manufacturers (2019).

36. A. Nazari, M. Jafari, N. Rezaei, F. Taghizadeh-Hesary, and F. Taghizadeh-Hesary, “Jet fans in the underground car parking areas and virus transmission.” Physics of Fluids 33.1, 013603 (2021).

37. D.N. Sørensen and P. v. Nielsen, “Quality control of computational fluid dynamics in indoor environments.” Indoor Air 13.1, 2–17 (2003).

38. A. Hasan, “Tracking the Flu Virus in a Room Mechanical Ventilation Using CFD Tools and Effective Disinfection of an HVAC System.” International Journal of Air - Conditioning and Refrigeration 28.2, 2050019 (2020).

39. H. Zhang, D. Li, L. Xie, and Y. Xiao, “Documentary Research of Human Respiratory Droplet Characteristics” Procedia Engineering 121, 1365–1374 (2015).

40. M. Vaish, “Lung-Aerosol Dynamics in Human Airway Models: Validation and Application of OpenFOAM Software” (2014).

41. J.Q. Feng, “A Computational Study of Particle Deposition Patterns from a Circular Laminar Jet” Journal of Applied Fluid Mechanics 10.4, 1001–1012 (2017).

42. Asadi, S., Wexler, A. S., Cappa, C. D., Barreda, S., Bouvier, N. M., & Ristenpart, W.D. “Aerosol emission and superemission during human speech increase with voice loudness.” Scientific Reports, 9.1, 1–10. (2019).

43. “Valedictorian^®^ - Vertical Unit Ventilator Models VFF and VFV” MODINE (2012)

44. J.K. Gupta, C.-H. Lin, and Q. Chen, “Characterizing exhaled airflow from breathing and talking” Indoor Air 20.1, (2010).

45. C. Pörschmann, T. Lübeck, and J.M. Arend, “Impact of face masks on voice radiation. “ The Journal of the Acoustical Society of America 148.6, 3663–3670 (2020).

46. G. Boiger, “Development and Verification of a Novel Lagrangian, (Non-)Spherical Dirt Particle and Deposition Model to Simulate Fluid Filtration Processes Using OpenFOAM”, (2009).

